# Neonatal methylation-based predictors of childhood cognition

**DOI:** 10.1101/2025.05.23.25328231

**Authors:** Rebekah Smikle, Katie Mckinnon, Kadi Vaher, Helen Turner, Hilary Cruickshank, Ray Amir, Yu Wei Chua, Eleanor LS Conole, Rebecca M Reynolds, G David Batty, Athanasios Tsanas, Lee Murphy, Heather C Whalley, Riccardo E Marioni, Simon R Cox, James P Boardman

**Affiliations:** Centre for Reproductive Health, Institute for Regeneration and Repair, University of Edinburgh, Edinburgh, UK; Centre for Clinical Brain Sciences, University of Edinburgh, Edinburgh, UK; School of Philosophy, Psychology, and Language Sciences, University of Edinburgh, Edinburgh, UK; Simpson Centre for Reproductive Health, Royal Infirmary Edinburgh, Edinburgh, UK; Department of Public Health, Policy and Systems, Institute of Population Health, University of Liverpool, Liverpool, UK; Department of Biochemistry, University of Oxford, Oxford, UK; Centre for Cardiovascular Science, University of Edinburgh, Edinburgh, UK; Department of Epidemiology and Public Health, University College London, London, UK; Usher Institute, Edinburgh Medical School, University of Edinburgh, Edinburgh, UK; The Alan Turing Institute, London, UK; Edinburgh Clinical Research Facility, University of Edinburgh, Edinburgh, UK; Centre for Genomic and Experimental Medicine, Institute of Genetics and Cancer, University of Edinburgh, Edinburgh, UK; Lothian Birth Cohorts, Department of Psychology, University of Edinburgh, Edinburgh, UK

## Abstract

Preterm birth is closely associated with immune dysregulation in early life and subsequent learning and psychiatric disorders, but methods for stratifying infants at risk remain elusive. Epigenetic Scores (EpiScores) are relatively stable DNA methylation (DNAm)-based proxies of circulating proteins that can capture health-related exposures such as chronic inflammation. EpiScore of C-reactive protein (DNAm CRP) is associated with inflammatory burden in early life, atypical brain development following preterm birth (encephalopathy of prematurity), and adult cognitive ability. To evaluate the utility of EpiScores for predicting cognition in children born preterm, we investigated relations between 43 neonatal saliva-based EpiScores known to associate with low gestational age, and cognition assessed at 2 and 5 years of age in a cohort of 232 preterm and term-born children.

DNAm CRP was negatively associated with 5-year Mullen Scales of Early Learning Composite (ELC) (β = −0.273, p = 0.002). Association magnitudes were larger for children born earlier (DNAm CRP x gestational age, β_interaction_ = 0.181). EpiScores of CRTAM, NCAM1 and SLITRK5 were also associated with 5-year ELC in the full cohort (absolute β range 0.219 to 0.267, Bonferroni-adjusted p-values <0.01). For preterm children, associations for DNAm CRP (β = −0.318, p = 0.021) and DNAm CRTAM (β = −0.307, p = 0.006) with 5-year ELC remained significant after adjustment for inflammatory exposures. We demonstrate associations between a range of neonatal salivary EpiScores and childhood cognition, suggesting the clinical value of EpiScores as early life markers of cognitive ability in children at risk of impairment warrants further investigation.

## Introduction

Mounting evidence in the field of developmental programming suggests that perinatal exposure to inflammation increases vulnerability to neurodevelopmental and psychiatric conditions, including autism, cerebral palsy, schizophrenia, depression and even Alzheimer’s disease (1,2). Despite the multifactorial and heterogeneous nature of these conditions, immune dysregulation in early life is considered a common pathological component of their aetiology.

Very preterm birth (before 37 completed weeks’ gestation) is often associated with a characteristic pro-inflammatory profile that arises in the context of multiple morbidities in the perinatal period compared to term infants (3–5). Elevation of selected pro-inflammatory blood proteins during the first postnatal month is associated with magnetic resonance imaging (MRI) measures of neonatal brain development (4) and cognition in several domains in early childhood (6–12). However, few studies include a comparator group of term-born controls, and there remains a need to identify early-life markers of immune dysregulation that have functional significance at school entry age, when cognitive demands increase and neuropsychological difficulties emerge (13). Moreover, measuring selected blood-based proteins at a single time point may not capture baseline and sustained inflammation, as their expression is often phasic (14,15), and serial blood sampling raises practical and ethical considerations in neonates. Consequently, methods for the early detection of children at risk of cognitive impairment due to immune dysregulation remain elusive.

Protein ‘Epigenetic scores’ (EpiScores) offer a novel, potentially more stable means of profiling inflammation and other biological processes compared to measuring circulating protein (16–19). EpiScores are DNA methylation (DNAm)-based biomarkers, derived from a linear weighted sum of DNAm at CpG sites, that correlate with protein expression levels (16,17). EpiScores are generated by identifying CpG sites relevant to particular traits or exposures through methods such as epigenome-wide association studies (EWAS) or penalised regression models; weights derived from these analyses are then applied to DNAm in an independent sample to compute EpiScores for that trait or exposure (20). In previous work we investigated their research utility when derived from neonatal saliva samples because preterm birth is associated with widespread changes in the salivary methylome (21), which we and others have shown is altered in response to inflammation in early life (21–23). Low gestational age at birth is associated with 43 out of 104 studied DNAm-based EpiScores enriched for inflammatory proteins (24). Notably, DNAm CRP correlates with low gestational age and perinatal inflammatory exposures (histologic chorioamnionitis, sepsis, bronchopulmonary dysplasia and necrotising enterocolitis), and is associated with MRI features of the encephalopathy of prematurity (EoP) (25).

Studies have reported associations between chronic, cumulative inflammation, indexed by EpiScores, and neurocognitive outcomes. For instance, multiple EpiScores associate with concurrent cognitive functioning in adult cohorts (18,26,27). Barker et al. (28) show that DNAm CRP in umbilical cord blood is associated with cognition at age 7 years; but other studies of EpiScores in paediatric populations have primarily focused on MRI features or mental health outcomes (29,30). In the context of preterm birth, specific CpG methylation patterns in placenta and buccal tissue have been associated with childhood cognition (31–33). However, epigenetic proxies of specific proteins, offering a non-invasive measure of perinatal inflammation, may be of translational utility for predicting brain-related health and cognitive outcomes in childhood.

In this study, we linked neonatal saliva-based EpiScores with measures of development and cognition at 2 and 5 years of age acquired from a birth cohort enriched for prematurity (34) to test four hypotheses. First, we tested a specifically directional hypothesis focusing on DNAm CRP given the comparatively rich prior literature for this marker: we hypothesised that higher DNAm CRP would be associated with poorer general cognitive ability at 2 and 5 years of age. Second, we investigated whether any of the 42 other EpiScores that are altered in preterm infants are associated with general cognitive ability at 2 and 5 years of age. Third, we examined whether there is an interaction between DNAm CRP and gestational age at birth, hypothesising that the association between DNAm CRP and general cognitive ability would be stronger for children born at lower gestational ages. Fourth, we tested the hypothesis that in preterm children, associations between EpiScores and general cognitive ability are attenuated by accounting for perinatal inflammatory exposures.

## Materials and Methods

### Participants

Participants were 2- and 5-year-old children born at <33 weeks’ gestation and a comparator group of children born at term. All participants were recruited at birth into the Theirworld Edinburgh Birth Cohort (TEBC), a prospective, longitudinal study designed to investigate the impact of preterm birth on brain development and outcomes (34). Cohort exclusion criteria were major congenital malformations, chromosomal abnormalities, congenital infection, parenchymal brain injury (cystic periventricular leukomalacia, haemorrhagic parenchymal infarction or post-haemorrhagic ventricular dilation). As such, the cohort is representative of the majority of survivors of modern neonatal intensive care practices (34,35). Ethical approval was obtained from the National Research Ethics Service, South East Scotland Research Ethics Committee (11/55/0061, 13/SS/0143 and 16/SS/0154). Informed consent was obtained from a person with parental responsibility for each participant. Neonatal DNAm data and cognitive assessment data at one or both time points (2 and 5 years) were available for 232 children.

### Clinical and demographic measures

Clinical and demographic data were collected through questionnaires administered to the primary caregiver and review of medical records. We included the following perinatal inflammatory comorbidities following definitions from previous work (4): histologic chorioamnionitis (HCA) was defined as the presence of inflammatory response in the placental membranes of any grade or stage; sepsis was defined as blood culture positive for pathogenic organism (culture positive) or clinically suspected infection treated with intravenous antibiotics for ≥5 days (culture negative); necrotising enterocolitis (NEC) was defined as medical treatment for ≥7 days or surgical treatment; bronchopulmonary dysplasia (BPD) was defined as the need for supplemental oxygen therapy or respiratory support at 36 weeks corrected gestational age. All comorbidities were coded as binary variables (present/absent). Birthweight z-scores were calculated according to INTERGROWTH-21st standards (36). Socioeconomic status was based on the Scottish Index of Multiple Deprivation 2016 (SIMD) and maternal education attainment (highest achieved) around the time of birth. SIMD, a neighbourhood-level indicator of socioeconomic circumstances, is derived from a family’s postal code. It uses multiple dimensions to assign a level of deprivation to the neighbourhood, ranked from most deprived (rank 1) to least deprived (rank 6976) (37). Maternal education, an individual-level indicator of socioeconomic status, was coded as a binary variable of a university undergraduate degree or higher (Yes/No). Maternal cigarette smoking during pregnancy (self-reported) was coded as current versus never/previous.

### DNA methylation and EpiScore calculation

Saliva samples for DNAm were obtained from preterm infants at term-equivalent age and from term infants shortly after birth. Saliva was collected in Oragene OG-575 Assisted Collection kits, by DNA Genotek, and DNA extracted using prepIT.L2P reagent (DNA Genotek, ON, Canada).

Details of methylation measurement have been described previously (24,25). Briefly, DNA was bisulphite converted and methylation levels were measured using Illumina HumanMethylationEPIC BeadChip (Illumina, San Diego, CA, USA) at the Edinburgh Clinical Research Facility (Edinburgh, UK). The arrays were imaged on the Illumina iScan or HiScan platform, and genotypes called automatically using GenomeStudio Analysis software 2011.1 (Illumina). DNAm was processed in four batches.

Details of DNAm data pre-processing have also been outlined previously (21,24). Raw intensity (.idat) files were read into the R environment (version 3.4.4) using *minfi. wateRmelon* and *minfi* were used for preprocessing, quality control, and normalisation (38,39). Specifically, the *pfilter* function in *wateRmelon* was used to exclude samples with 1% of sites with a detection p-value > 0.05, sites with beadcount < 3 in 5% of samples, and sites with 1% of samples with detection p-value > 0.05. Cross-hybridising probes, probes targeting single nucleotide polymorphisms with overall minor allele frequency ≥0.05 and control probes were also removed (40). Samples were removed if there was a mismatch between predicted sex (*minfi*) and observed sex (n = 3), or if samples did not meet preprocessing quality control criteria (n = 29). Data were danet normalised which includes background correction and dye bias correction (38). Saliva contains different cells types, including buccal epithelial cells. Epithelial cell proportions were estimated with epigenetic dissection of intra-sample heterogeneity with the reduced partial correlation method implemented in the R package EpiDISH (41). Probes located on sex chromosomes were removed before analysis. For each participant, EpiScores were obtained by multiplying the methylation proportion at a given CpG by the effect size from previous studies, as previously described (16,24).

### Cognitive assessments

#### Bayley Scales of Infant and Toddler Development

At around 24 months corrected age, preterm children born <1500 g or <32 weeks of gestation were assessed using the Bayley Scales of Infant and Toddler Development, Third Edition (Bayley-III) (42) by trained practitioners blinded to EpiScores. The Bayley-III cognitive composite provides a measure of general cognitive ability across memory, information processing, concept formation, problem solving and categorisation; it is an age-standardised index score, with normative mean of 100 and standard deviation (SD) of 15. One hundred and fifty-four participants with neonatal EpiScore data had cognitive data at the 2-year time-point. Their data corresponded to DNAm processing batches 1-4: batch 1 included 74 participants (48.1%); batch 2 included 40 (26.0%); batch 3 included 21 (13.6%); batch 4 included 19 (12.3%).

#### Mullen Scales of Early Learning

At around 5 years of age, children were assessed using the Mullen Scales of Early Learning (MSEL) (43). The MSEL Early Learning Composite (ELC) provides a measure of general cognitive performance, derived from four subscales: visual reception, fine motor, receptive language and expressive language. ELC is a standard score with normative mean of 100 and SD of 15. In exploratory analyses defined below, we also derived the age-adjusted subscale scores, which are each T*-* scores with mean of 50 and SD of 10. One hundred and twenty-seven participants with neonatal EpiScore data had MSEL ELC data at 5 years of age. Their data corresponded to DNAm processing batches 1 and 2: batch 1 included 50 participants (39.4%); batch 2 included 77 (60.6%).

### Statistical analysis

The analysis plan for this project was preregistered (https://osf.io/pghw2) and all statistical analyses were performed in R (version 4.2.3).

Maternal and infant clinical and demographic characteristics were compared between preterm and term children. Standardised mean difference (Cohen’s d) was used to quantify effect sizes: 0.2 (small), 0.5 (medium), and 0.8 (large) (44). For continuous variables, we used Mann-Whitney *U*-test to compare distributions. For categorical variables, we used Chi-square tests to compare proportions.

All regression analyses in this study included baseline and adjusted models. Outcomes of interest were Bayley-III Cognitive Composite scores or MSEL Early ELC scores. Covariates for each model are specified below. Covariates were selected based on their potential to act as confounders—variables that influence both EpiScores and the outcome—or because their inclusion could improve the precision of the model estimates. Adjusted models accounted for prematurity-related and socioeconomic variables. Continuous variables were scaled to have a mean of zero and variance of one. In the event of missing data for any covariates, complete case analyses were performed for comparison between baseline and adjusted multivariable models.

Baseline models: outcome of interest ∼ EpiScore + gestational age at sampling + sex + batch

Adjusted models: outcome of interest ∼ EpiScore + gestational age at sampling + sex + batch + gestational age at birth + birthweight Z-score + maternal education + SIMD

#### DNAm CRP and cognitive ability associations

To determine the effect of neonatal DNAm CRP on Bayley-III Cognitive composite at 2 years or MSEL ELC at 5 years, data were analysed in univariable, baseline and adjusted regression models. Visual inspection of diagnostic plots suggested no regression assumptions were violated (Supplementary Figure 1). Analyses were performed for whole group, and also stratified by preterm or term children. Given that an interaction effect has been observed between DNAm CRP and gestational age at birth on MRI metrics, such that there were significant relationships found in preterm but not term neonates (25), we repeated analyses to include a product interaction term (DNAm CRP x gestational age at birth), for the whole group and for the preterm subgroup.

In models where DNAm CRP was the predictor variable, we conducted one-tailed significance tests given the specific direction of our preregistered hypothesis, and thus applied a p-value significance threshold of <0.10. This preregistered threshold was applied to DNAm CRP because of its known associations with GA, perinatal inflammatory burden and MRI features of EoP (25).

#### DNAm CRP and MSEL subscale associations

In exploratory analyses, we performed linear regressions to examine associations between DNAm CRP and MSEL subscales in univariable, baseline and adjusted models. Here, we corrected p-values for multiple comparisons using the false discovery rate (FDR) method (Benjamini-Hochberg procedure (45)), at the level of MSEL subscales in each model type separately. Significance was deemed at a threshold of p_FDR_ (*q-*value) <0.05. We further compared the magnitude of associations across domains and determined the relative effects of DNAm CRP on specific cognitive domains. Differences between association magnitudes were assessed using the Williams (1959) (46) test for dependent groups with overlapping correlations, using the “cocor” R package (http://cran.r-project.org/web/packages/cocor/cocor.pdf).

#### Other EpiScore and cognitive ability associations

To determine the associations of the other 42 neonatal EpiScores with Bayley-III Cognitive composite at 2 years or MSEL ELC at 5 years, data were similarly analysed in univariable, baseline and adjusted regression models. Analyses were performed for whole group and preterm subgroup. For these models with the 42 EpiScores, we first estimated the number of independent statistical tests by conducting a principal component analysis (PCA). This approach, consistent with our previous work (24), accounts for the high intercorrelation among EpiScores by reducing dimensionality. The resulting estimate of statistical tests allowed us to apply a Bonferroni correction for multiple comparisons that was appropriately stringent without being overly conservative. Spearman’s correlation analysis of EpiScores showed (absolute) correlation coefficients of 4.71 x 10^-5^ to 0.927 (Figure 1A). PCA using Kaiser Criterion yielded ten principal components with eigenvalues >1 from the 42 EpiScores (Figure 1B; Supplementary Table 1), explaining 69.5% cumulative variance. Inspection of the scree plot (Figure 1C) revealed an elbow at the fifth principal component, explaining 55.0% cumulative variance. Thus, our analyses report associations which survive correction for multiple comparisons using two Bonferroni-adjusted significance thresholds: a stricter threshold of p <0.005 (0.05/10), corresponding to the ten principal components identified using Kaiser Criterion, and a less stringent threshold of p <0.01 (0.05/5), corresponding to the five principal components identified using the elbow method.

**Figure 1.**
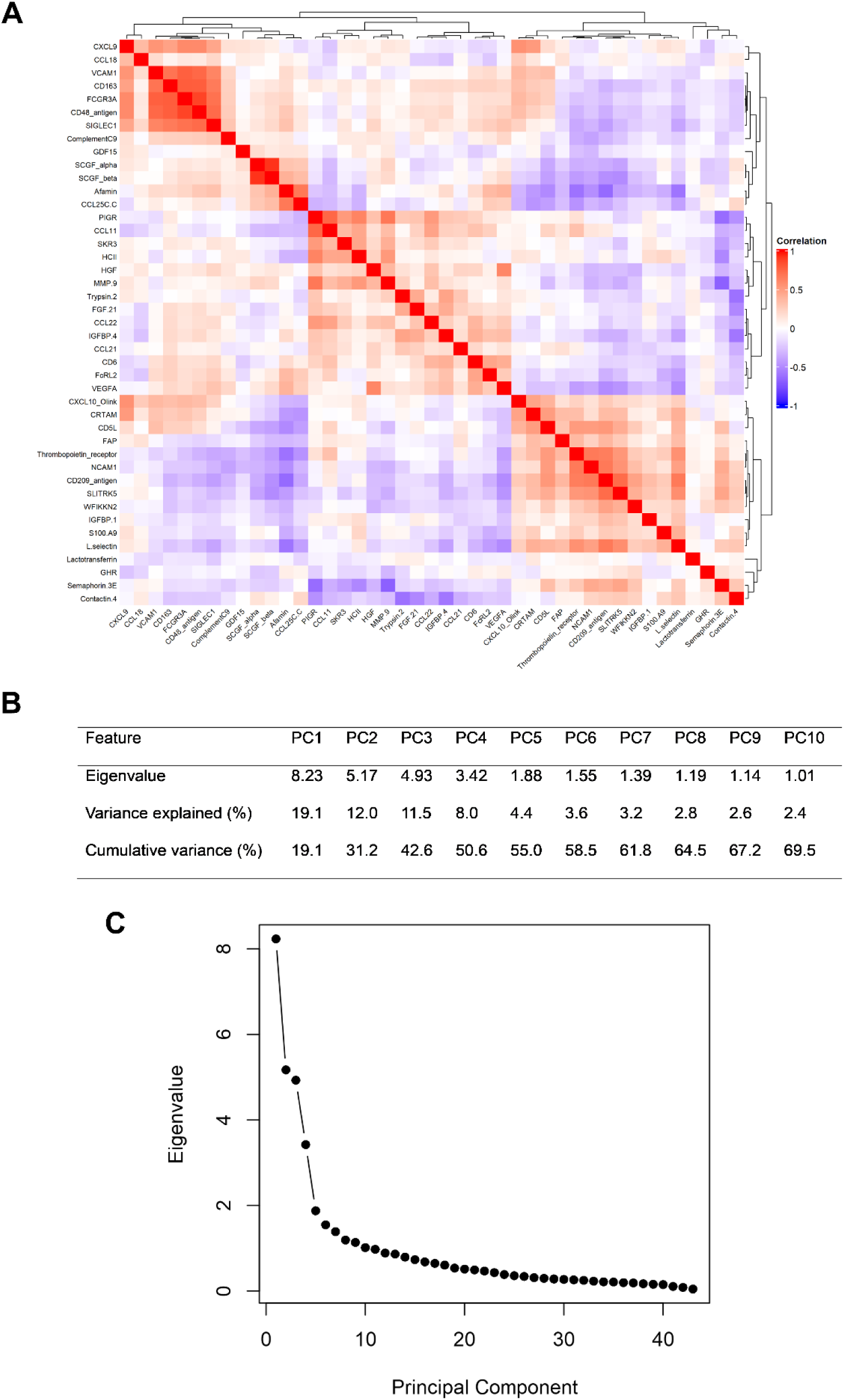
Dimensionality reduction of EpiScores to determine statistical families for correction of multiple comparisons. A) Heatmap and dendrogram of correlations of EpiScores. Direction and magnitude of Spearman’s correlation coefficients are shown by the colour scale, with red as positive and blue as negative when statistically significant (p <0.05). B) Principal components analysis eigenvalues and variance explained by each principal component. C) Scree plot of eigenvalues against principal components of EpiScores. PC = principal component.

#### EpiScore and cognitive ability associations, accounting for perinatal inflammatory exposures

Finally, to assess the role of inflammation in explaining the associations, we conducted subgroup analyses in preterm children, additionally accounting for the total number of perinatal inflammatory morbidities (histological chorioamnionitis, sepsis, BPD and NEC). The cumulative burden of perinatal inflammation – indexed by total number of these inflammatory exposures – was previously shown to be positively associated with DNAm CRP (25). In baseline and adjusted models where inflammatory exposures were included as covariates, we quantified the degree of attenuation in the main effect of DNAm CRP or other EpiScores by comparing magnitude of the hierarchical standardised regression coefficients (β). To quantify the variance in MSEL ELC explained by DNAm CRP or other EpiScores beyond that explained by inflammatory exposures, we also reported both the adjusted R^2^ and incremental R^2^. Incremental R^2^ was calculated by comparing the R^2^ of each model to that of a null model in which MSEL ELC was modelled with only sex, gestational age at sampling and batch.

## Results

### Participant characteristics

At 2 years, n = 154 preterm children with neonatal EpiScores were assessed for Bayley-III, characteristics of whom are reported in Supplementary Table 2. At 5 years, 127 children (67 preterm) were assessed for MSEL, characteristics of whom are shown in Table 1. MSEL ELC scores tended to be lower in preterm children (Table 1).

**Table 1.** Demographic and clinical characteristics of participants with neonatal EpiScores and 5-year Mullen Scales of Early Learning (MSEL) data (n = 127)

|  | Term | Preterm | SMD |
| --- | --- | --- | --- |
| n | 60 | 67 |  |
| Sex: Female, n (%) | 24 (40.0%) | 31 (46.3%) | 0.127 |
| Gestational age at birth / weeks, median (range) | 39.64 (36.43, 42.14) | 29.42 (25.14, 32.00) | 6.796 |
| Gestational age at DNAm sampling / weeks, median (range) | 42.14 (40.00, 46.14) | 40.57 (37.70, 43.43) | 1.324 |
| Neonatal DNAm CRP, median (range) | -0.011 (-0.012, -0.010) | -0.010 (-0.013, -0.010) | 0.292 |
| Chronological age at MSEL assessment / months, median (range) | 60.5 (59.0, 63.9) | 60.4 (59.4, 65.3) | 0.152 |
| MSEL Early Learning Composite, median (range) | 97 (70, 121) | 89 (60, 127) | 0.349 |
| Birthweight / g, median (range) | 3454 (2410, 4560) | 1112 (600, 2100) | 6.264 |
| Birthweight z-score, median (range) | 0.45 (-2.30, 2.57) | -0.03 (-2.62, 1.58) | 0.698 |
| Histological chorioamnionitis, n (%) | 3 (8.6) | 16 (24.6) | 0.442 |
| Sepsis, n (%) | NA | 14 (20.9) | - |
| Bronchopulmonary dysplasia, n (%) | NA | 21 (32.3) | - |
| Necrotising enterocolitis, n (%) | NA | 2 (3.0) | - |
| Maternal age / years, median (range) | 33 (24, 47) | 33 (20, 44) | 0.283 |
| Maternal BMI / median (range) | 23.90 (18.20, 47.00) | 25.10 (16.40, 48.00) | 0.185 |
| Smoked during pregnancy, n (%) | 0 (0.0) | 6 (9.1) | 0.447 |
| Corticosteroid administration in pregnancy, n (%) | 0 (0.0) | 64 (95.5) | 6.532 |
| MgSO<sub>4</sub> administration in pregnancy, n (%) | NA | 50 (74.6) | - |
| Mother university degree, n (%) | 54 (90.0) | 42 (62.7) | 0.679 |
| SIMD rank, median (range) | 5471 (727, 6951) | 4475 (137, 6929) | 0.461 |

For categorical data, Chi-square test was used to compare proportions. For continuous data, Mann-Whitney *U*-test was used. Standardised mean difference, SMD, is derived from Cohen's *d* for continuous data. Values are denoted as n (%) or median (range). SIMD = Scottish Index of Multiple Deprivation.

### Associations between DNAm CRP and general cognitive abilities

At 5 years, median neonatal DNAm CRP was significantly higher in preterm compared to term-born children (SMD = 0.292; Table 1; Figure 2A). Higher DNAm CRP was associated with lower MSEL ELC scores (5 years) in whole group analyses (n = 125) in univariable models and baseline models (β = −0.273 and −0.165, respectively, p <0.10), consistent with the directional hypothesis (Figure 2B). However, magnitude of associations between DNAm CRP and MSEL ELC were reduced to non-significance after adjusting for gestational age at birth and socioeconomic exposures (absolute percentage decrease in β = 3%; p = 0.157; Figure 2B), suggesting Simpson’s paradox where gestational age may be either a confounder or an interaction term (47). Indeed, whole group analyses revealed significant interaction effects between gestational age and DNAm CRP in baseline models (product interaction term β = 0.181; main effect β = −0.034) and adjusted models (product interaction term β = 0.240; main effect β = −0.064), indicating that at lower gestational ages, the negative effect of DNAm CRP on MSEL ELC is stronger (Figure 3; Supplementary Table 5).

**Figure 2.**
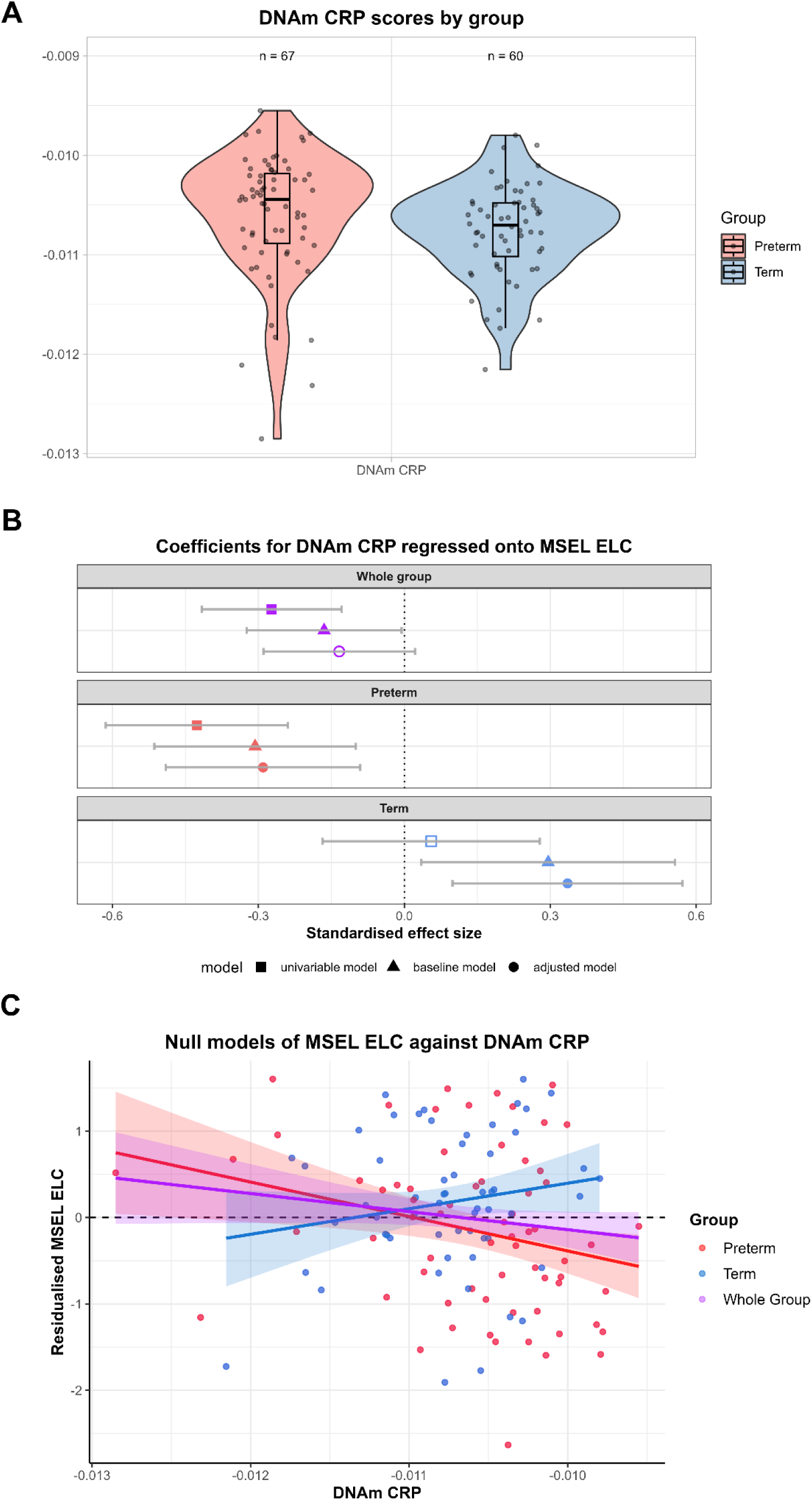
Neonatal DNAm CRP in preterm and term-born children and its association with MSEL Early Learning Composite (ELC), illustrating Simpson’s paradox. A) Violin and box plots of DNAm CRP by group. B) Forest plots for DNAm CRP associations with MSEL ELC for whole group, preterm and term-born children. Points show standardised regression coefficients and 90% confidence intervals (tests of specific, directional, pre-registered hypotheses). Significant associations are denoted by filled shapes, where p <0.10. Baseline models are controlled for infant sex, gestational age at sampling and batch. Adjusted models additionally control for gestational age at birth, birthweight Z-score, maternal university education and SIMD. C) Scatterplots with regression lines by gestational age, conveying Simpson’s paradox. Residuals from null models (adjusted for infant sex, gestational age at sampling and batch) are plotted against DNAm CRP. MSEL = Mullen Scales of Early learning, SIMD = Scottish Index for Multiple Deprivation.

**Figure 3.**
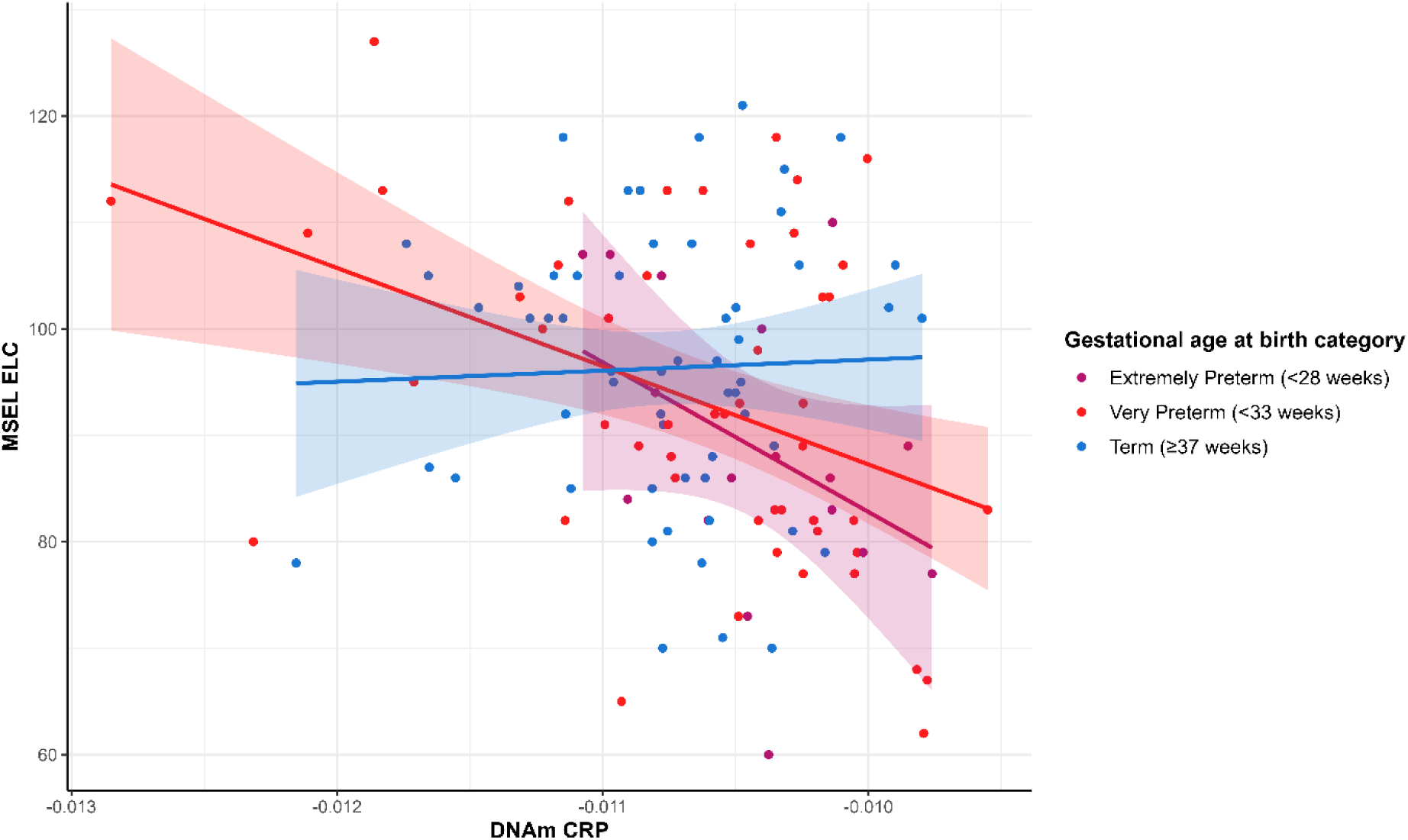
Interaction effects of DNAm CRP and gestational age on MSEL Early Learning Composite (ELC) at five years of age. Interactions are illustrated as three groups for ease of interpretation, namely extremely preterm, very preterm and term.

The interaction effect was further supported by subgroup analyses (Figure 2B; Supplementary Table 4). Specifically, in the preterm group (n = 67), higher DNAm CRP was consistently associated with lower MSEL ELC scores across all models (βs = −0.290 to −0.427, p <0.10). In contrast, among term-born children (n = 58), univariable models showed no association (β = 0.055, p = 0.680), whereas inclusion of covariates in baseline and adjusted models revealed a positive association (β = 0.296 and 0.335, respectively; p <0.10). This positive association appeared to be driven by adjustment for processing batch (batch β = −1.167, p = 0.001) as no other covariates showed significant associations; notably, batch was not correlated with other covariates suggesting it acted as a suppressor variable.

DNAm CRP was not associated with 2-year Bayley-III Cognitive composite in univariable analyses or in baseline models adjusted for gestational age at sampling, sex and processing batch, or models adjusted for additional prematurity-related and socioeconomic variables (all β’s ≤ |0.024|, p >0.05; Supplementary Table 3).

### Associations between DNAm CRP and domain-specific cognitive abilities

Given that DNAm CRP was associated with global cognitive ability at 5 years, we investigated whether associations are domain-specific using MSEL subscales (Supplementary Figure 2). Higher DNAm CRP was associated with lower MSEL score across most subscales in univariable analyses for the whole group (β = −0.203 to −0.243, p_FDR_ <0.05; Supplementary Table 6) and preterm subgroup (β = −0.258 to - 0.485, p_FDR_ <0.05; Supplementary Table 7). Further, associations in preterm children remained significant for fine motor (β = −0.284 to −0.314, p_FDR_ <0.05) and visual reception (β = −0.396 to −0.437, p_FDR_ = <0.05) in baseline and adjusted models. Comparing magnitudes of DNAm CRP associations across domains in preterm baseline models revealed significant differences between visual reception and both receptive language (t = −3.143, p =0.002) and expressive language (t = −2.602, p = 0.009); no differences were observed between other domains (Supplementary Table 8).

### Associations between other EpiScores and general cognitive abilities

At 5 years, DNAm SLITRK5 (β = 0.242) and DNAm CD209 (β = 0.267) were positively associated with MSEL ELC in univariable analyses, with the latter also meeting the more stringent Bonferroni-adjusted p <0.005. Five other EpiScores were negatively (β = −0.178 to −0.228) and three positively associated (β = 0.192 to 0.228) with 5-year MSEL ELC scores, but these nominal associations did not meet Bonferroni-adjusted p <0.01 (Figure 4; Supplementary Table 10). In baseline models covarying for gestational age at sampling, sex and processing batch, higher DNAm CRTAM was significantly associated with poorer MSEL ELC score (β = −0.219 [95% CI −0.384, −0.054], Bonferroni adjusted p <0.01). DNAm HCII (β = −0.245) and DNAm SCGF alpha (β = 0.197) were also nominally associated with MSEL ELC at the uncorrected threshold (all p <0.05). No EpiScore associations survived additional adjustment for prematurity-related or socioeconomic variables in whole group analyses.

**Figure 4.**
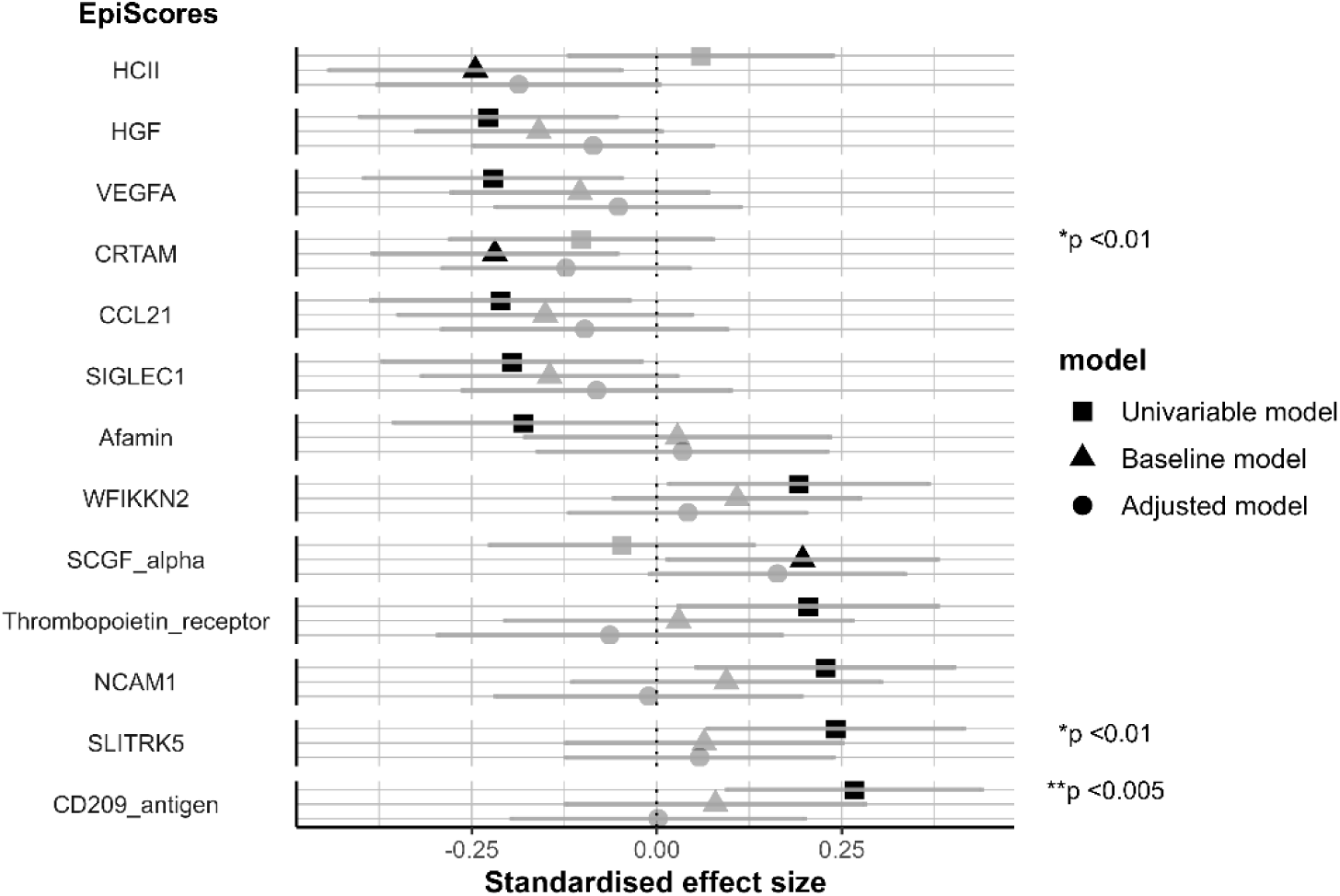
Association between EpiScores with MSEL Early Learning Composite. Points show standardised regression coefficients and 95% confidence intervals. Significant associations at the nominal threshold p <0.05 are denoted by black shapes. Asterisks indicate significant associations at Bonferroni-adjusted thresholds: (*) p <0.01 and (**) p <0.005. Baseline models are controlled for infant sex, gestational age at sampling and batch. Adjusted models additionally control for gestational age at birth, birthweight Z-score, maternal university education and SIMD. Models showing significant associations are ordered by magnitude. MSEL = Mullen Scales of Early learning, SIMD = Scottish Index for Multiple Deprivation.

DNAm CCL18 (β = 0.198 [95% CI 0.036 to 0.361], p = 0.017) and DNAm CD5L (β = 0.238 [95% CI 0.016 to 0.460], p = 0.036) were nominally associated with 2-year Bayley-III Cognitive composite in baseline models. However, associations did not survive Bonferroni correction (adjusted p <0.01) or adjustment for other variables (Supplementary Table 9).

### Associations between EpiScores and cognition, accounting for perinatal inflammatory exposures in preterm infants

Finally, we tested the hypothesis that for preterm children exposure to perinatal inflammatory events partly accounts for the association between EpiScores and poorer cognitive outcomes at 5 years. Main effect size of association between DNAm CRP and MSEL ELC remained statistically significant and unattenuated after controlling for perinatal inflammatory exposures in preterm children (β = −0.318, absolute percentage β increase = 1.1%, p = 0.021; Table 2). Calculating the difference in R^2^ for baseline model and model after adjusting for inflammatory exposures showed a marginal increase of 0.1% in explained R^2^.

**Table 2.** Associations between DNAm CRP and MSEL Early Learning Composite, adjusting for number of inflammatory exposures in preterm children only.

| model | Beta | Lower CI | Upper CI | p | R<sup>2</sup> | Incremental R<sup>2</sup> | n |
| --- | --- | --- | --- | --- | --- | --- | --- |
| baseline model | **-0.307** | -0.514 | -0.100 | **0.016** | 0.263 | 0.060 | 67 |
| baseline model +<br>inflammatory<br>exposures | **-0.318** | -0.543 | -0.093 | **0.021** | 0.264 | 0.061 | 65 |
| adjusted model +<br>inflammatory<br>exposures | **-0.323** | -0.532 | -0.113 | **0.013** | 0.400 | 0.197 | 63 |

Betas (standardised regression coefficients for DNAm CRP) and 90% confidence intervals are reported. Incremental R<sup>2</sup> refers to the amount of variance in MSEL ELC accounted for by DNAm (and other inflammatory, prematurity-related or socioeconomic exposures), beyond that of baseline covariates: sex, gestational age at sampling and batch. Bold text denotes p<0.10.
Baseline model = MSEL ELC ~ DNAm CRP + sex + gestational age at sampling + batch
Adjusted model = MSEL ELC ~ DNAm CRP + sex + gestational age at sampling + batch + gestational age at birth + birthweight Z score + SIMD + maternal university education

DNAm CRTAM was also negatively associated with MSEL ELC (β = −0.307 [95% CI - 0.523, −0.090], Bonferroni-adjusted p = 0.006) in preterm subgroup, which remained significant after adjustment for inflammatory exposures (absolute percentage change in β = 0%; Bonferroni-adjusted p = 0.008; Supplementary Table 11). Models further adjusted for prematurity-related and socioeconomic variables retained nominal significance (β = −0.265, p = 0.016), but no longer met the Bonferroni-adjusted p <0.01.

## Discussion

We leveraged summary statistics from large-scale EWAS and penalised regression models to calculate methylation-based biomarkers, EpiScores, in neonatal saliva and integrated these with cognitive outcome data at two timepoints in childhood to investigate relations between chronic low-grade systemic perinatal inflammation and later cognition. Our key, novel finding was that four neonatal EpiScores are significantly associated with general cognition at age 5 years, with an additional twelve demonstrating nominal (uncorrected) significance at either 2 or 5 years. We showed that higher neonatal DNAm CRP associated with poorer 5-year MSEL Early Learning Composite, with moderate effect size, β = −0.163 to −0.307, the strength of which is more pronounced with decreasing gestational age at birth. Associations were independent of the number of inflammatory exposures, suggesting unique contributions of specific immuno-epigenetic signatures to variance in cognition beyond immune-related morbidities.

Given previously established associations of neonatal DNAm CRP with gestational age at birth, perinatal inflammatory exposures and MRI features of EoP (24,25), we examined the relation of this marker with cognition in this study separately to the other EpiScores which are associated with gestational age at birth. The observation that higher neonatal DNAm CRP associates with lower cognitive score in 5-year-olds is consistent with findings that blood-derived inflammation-related epigenetic polygenic risk scores are associated with childhood cognition (28). Here, we show CRP EpiScore derived using non-invasive sampling methods is associated with cognition at school age, and that the magnitude of effect is dose-dependent with degree of prematurity. This finding aligns with previous observations of elevated inflammatory protein concentrations, including CRP, in early life associating with cognition in extremely preterm infants (6,48), lending support to the clinical value of this methylation-based proxy. The analogous protein is an acute-phase reactant in the immune response, but the mechanisms through which it may impact cognition remain unclear. Notably, DNAm CRP is not associated with 2-year cognition according to the Bayley-III, which could be a function of test performance leading to overestimation of cognitive ability (49,50). Alternatively, DNAm CRP and most other EpiScores may not be sufficiently sensitive to detect differences in earlier stages of cognitive development.

Follow-up analyses with 5-year MSEL subscales revealed negative associations between DNAm CRP and all domains, with magnitudes largest for fine motor and visual reception in preterm children, even after adjusting for prematurity-related and socioeconomic factors. Williams test (46) confirmed that visual reception was disproportionately associated with DNAm CRP compared to language domains. The visual reception MSEL subscale assesses visual information processing, which depends on unimpaired development of ocular and visual cognitive functions. Whereas previous studies demonstrate associations between serum CRP and development of retinopathy of prematurity (ROP) (51,52), a hallmark preterm morbidity leading to visual impairment, our previous work showed no independent association between DNAm CRP at term-equivalent age and higher odds of requiring treatment for ROP in this sample (25). An alternative explanation is that neonatal DNAm CRP is associated with cerebral visual impairment, which is common among preterm-born school children and thought to be related to weaker structural connectivity in visual networks secondary to diffuse white matter dysmaturation (53). Thus, the relation between inflammatory burden of prematurity captured by neonatal DNAm CRP and cerebral visual function warrants further investigation, particularly since visual processing is important for development of language, motor and other skills in early life.

In addition to DNAm CRP, we found evidence that other EpiScores may be associated with childhood cognition. Though these other EpiScores have not been previously studied in a neurodevelopmental context, DNAm NCAM1 and DNAm SLITRK5, positively associated with 5-year cognition in this study, were associated with slower time-to-dementia in an adult cohort (27). Their analogous proteins, which are localised at synaptic sites (54,55), have been implicated in cognition-related biological processes. NCAM1 is a neuronal and glial cell adhesion molecule, which supports neuronal growth and synaptic plasticity. It has been implicated in neurocognitive impairment in Alzheimer’s disease and psychiatric disorders (56). SLITRK5 is a transmembrane protein widely expressed in neural tissue that supports neurogenesis, and is a suggested therapeutic target in several nervous system conditions (57). Overall, several of the analogous proteins associated with 5-year cognition are involved in the immune response (HCII, CRTAM), vascular development (VEGFA), or are growth factors (HGF, SIGLEC1, WFIKKN2, SCGF alpha), chemokines (CCL21) or cell membrane proteins (Thrombopoietin receptor, CD209, Afamin). VEGFA, in particular, was previously indicated in the extremely low gestational age newborn cohort to associate with neurodevelopmental outcomes (6). In respect to the two EpiScores positively associated with 2-year cognition in the preterm group, low cord blood levels of the chemokine CCL18 have previously been linked to an increased risk of cerebral palsy in preterm children (58), while plasma CD5L – implicated in both protective and deleterious inflammatory processes – has been shown to positively associate with dementia risk (59). Although majority of associations did not survive adjustment for multiple comparisons and other exposures in this moderately-sized sample, they may still warrant further investigation.

While most EpiScore-cognition associations did not remain statistically significant when analyses were repeated in the 5-year preterm subgroup, DNAm CRP and DNAm CRTAM associations persisted after accounting for inflammatory exposures. Indeed, accounting for morbidities for both proxies did not considerably increase explained variance of either model, suggesting a unique, more substantial contribution of the EpiScores over residual inflammatory experience. This is consistent with our previous study (25) showing similarly robust, independent associations of DNAm CRP with white matter microstructure, and highlights CRTAM as another notable target biomarker in this population. While not previously studied in the context of preterm birth, CRTAM has been highlighted as an immune-related Alzheimer’s disease risk gene using post mortem tissue (60).

Many EpiScores in this study were previously found to associate with MRI-derived brain volumes in adulthood, both in cognitively normal and at-risk adult cohorts (26,27). Various MRI metrics also partly mediated DNAm CRP-cognition association, with white matter volume contributing most strongly (26). We previously showed that in preterm neonates, DNAm CRP associated with regional and global brain volumes, and most strongly with white matter microstructure (25). In a predominantly term cohort, DNAm CRP derived in cord blood has similarly been associated with white matter development (30). It may be that the particular vulnerability in preterm children to widespread brain alterations is augmented by inflammatory experience, which in turn contributes to poorer childhood cognition (61). Sustained elevation of inflammatory proteins in extremely preterm infants was demonstrated to associate with lower IQ at age 10, even after adjusting for various brain volume metrics (8). In another study, inflammatory events predicted alterations in corpus callosum microstructure, which correlated with cognitive performance at age 6 (62). Given that EpiScores outperform proteomic associations with brain structure (26,63) and are associated with childhood cognition, future work could investigate whether white matter microstructure mediates EpiScore-cognition associations. Several EpiScores identified here may be indicative of disrupted neurodevelopmental processes including neurogenesis, synaptogenesis and myelination (13,61).

Strengths of this study include the range of multimodal data allowing us to profile numerous epigenetic biomarkers and characterise cognition at multiple time points, as well as account for relevant inflammatory, prematurity-related and socioeconomic factors. Particularly in this population, use of salivary-based methylation proxies of inflammatory proteins offers a non-invasive, more stable, attractive alternative to blood sampling in neonates. The shared ectodermal origins of buccal and neural tissue as well as high correlation of DNA methylation patterns in both tissues (*R* = 0.85) support the use of salivary-based EpiScores for investigations regarding cognition (64). The correlation of EpiScores with gestational age and DNAm CRP with perinatal morbidities indicates clinical salience in this distinct population (24,25). While the prospective nature of our data offers the strength of longitudinal observations, our preterm-term subgroup analyses in particular may be limited by the modest sample size. Finally, approaches such as Mendelian randomisation and preclinical studies are necessary to determine causality and isolate confounding genetic and environmental factors. Of interest would be the stability of these associations, given the sustained inflammation experienced by preterm infants (65). Nonetheless, the present study provides essential discovery insights motivating work towards individual-level prediction, validation and replication in prognostic studies, where positive and negative predictive values of neonatal EpiScores for cognition can be estimated.

Overall, in a cohort of children spanning the whole gestational age range, we found that neonatal methylation-based EpiScores were associated with general cognition at 5 years. In preterm children, associations were independent of perinatal inflammatory exposures. Our findings point to potential translational utility of peripheral methylation-based biomarkers for stratifying infants at risk of cognitive difficulties at preschool age.

## Data Availability

Anonymised data including EpiScores used in these analyses are deposited in Edinburgh DataVault (https://doi.org/10.7488/e65499db-2263-4d3c-9335-55ae6d49af2b) (66) and are available to researchers under the terms of the Theirworld Edinburgh Birth Cohort Data Access and Collaboration policy available at https://reproductive-health.ed.ac.uk/theirworld-edinburgh-birth-cohort-tebc/for-researchers/data-access-and-collaboration.

## Acknowledgements

The authors are grateful to the families of the TEBC who consented to take part in the study. We thank Selina Abel and Melissa Thye for substantial contributions to MSEL data collection.

## CRediT author contributions

RS: conceptualisation, methodology, formal analysis, investigation, resources, data curation, writing – original draft, writing – review & editing, visualisation, funding acquisition.

KM: methodology, investigation, resources, data curation, writing – review & editing.

KV: investigation, methodology, data curation, writing – review & editing, visualisation.

HT: investigation, resources, data curation, writing – review & editing.

HC: investigation, resources, data curation, project administration, writing – review & editing.

RA: investigation, resources, data curation, project administration, writing – review & editing.

YWC: investigation, resources, data curation, project administration, writing – review & editing.

ELSC: methodology, data curation, writing – review & editing, visualisation.

RMR: investigation, resources, writing – review & editing.

GDB: investigation, resources, writing – review & editing.

AT: investigation, resources, writing – review & editing.

LM: investigation, resources, writing – review & editing.

HCW: methodology, writing – review & editing, funding acquisition.

REM: methodology, resources, writing – review & editing, funding acquisition.

SRC: conceptualisation, methodology, formal analysis, investigation, resources, data curation, writing – original draft, writing – review & editing, visualisation, supervision, funding acquisition.

JPB: conceptualisation, methodology, formal analysis, investigation, resources, data curation, writing – original draft, writing – review & editing, visualisation, supervision, project administration, funding acquisition.

## Ethics Declarations Conflict of Interest

LM has received speaker and consultancy fees from Illumina. REM is a scientific advisor to the Epigenetic Clock Development Foundation and to Optima partners. All other authors declare that they have no competing interests.

## Data availability

Anonymised data including EpiScores used in these analyses are deposited in Edinburgh DataVault (https://doi.org/10.7488/e65499db-2263-4d3c-9335-55ae6d49af2b) (66) and are available to researchers under the terms of the Theirworld Edinburgh Birth Cohort Data Access and Collaboration policy available at https://reproductive-health.ed.ac.uk/theirworld-edinburgh-birth-cohort-tebc/for-researchers/data-access-and-collaboration. Source code for primary analysis and figures are available at https://git.ecdf.ed.ac.uk/jbrl/neonatal-episcores-and-childhood-cognition.

