## Supplementary material for "Neonatal methylation-based predictors of childhood cognition": Supplemetary Information

Affiliations

**This document includes**

- Supplementary Figure 1. Model diagnostic metrics for association of DNAm CRP with MSEL Early Learning Composite. Assumptions shown are tested for baseline model = MSEL ELC ~ DNAm CRP + sex + GA sampling + batch
- Supplementary Table 1. Loadings of the 42 EpiScores onto the first 10 principal components.
- Supplementary Table 2. Demographic and clinical characteristics of participants with neonatal EpiScores and 2-year Bayley Scales of Infant and Toddler Development (Bayley-III) data.
- Supplementary Table 3. Association of DNAm CRP with Bayley-III Cognitive Composite in preterm children at 2 years old.
- Supplementary Table 4. Association of DNAm CRP with MSEL Early Learning Composite (ELC) in preterm children and term-born children at 5 years old.

- Supplementary Table 5. Interaction effects between DNAm CRP and gestational age on MSEL Early Learning Composite in full cohort and preterm only. Standardised betas and p-values are reported from regression models where DNAm CRP is regressed onto MSEL ELC, with and without an interaction term, DNAm:CRP. Bold text denotes p <0.10.
- Supplementary Figure 2. Associations between DNAm CRP and MSEL subscales in whole group and preterm subgroup.
- Supplementary Table 6. Associations between DNAm CRP and MSEL subscales in exploratory analyses in the full cohort at 5 years old (n = 125).
- Supplementary Table 7. Associations between DNAm CRP and MSEL subscales in exploratory analyses in the preterm children only at 5 years old (n = 67).
- Supplementary Table 8**.** Williams' test (1959) for differences in magnitudes of standardised DNAm CRP regression coefficients for baseline models between MSEL subscales: fine motor (FM), visual reception (VR), expressive language (EL), receptive language (RL).
- Supplementary Table 9. Association of EpiScores with Bayley-III Cognitive Composite in preterm children at 2 years old (n = 150).
- Supplementary Table 10. Association of EpiScores with MSEL Early Learning Composite (ELC) in the full cohort at 5 years old (n = 125).
- Supplementary Table 11. Associations between EpiScores and MSEL Early Learning Composite, adjusting for number of inflammatory exposures in preterm children only.

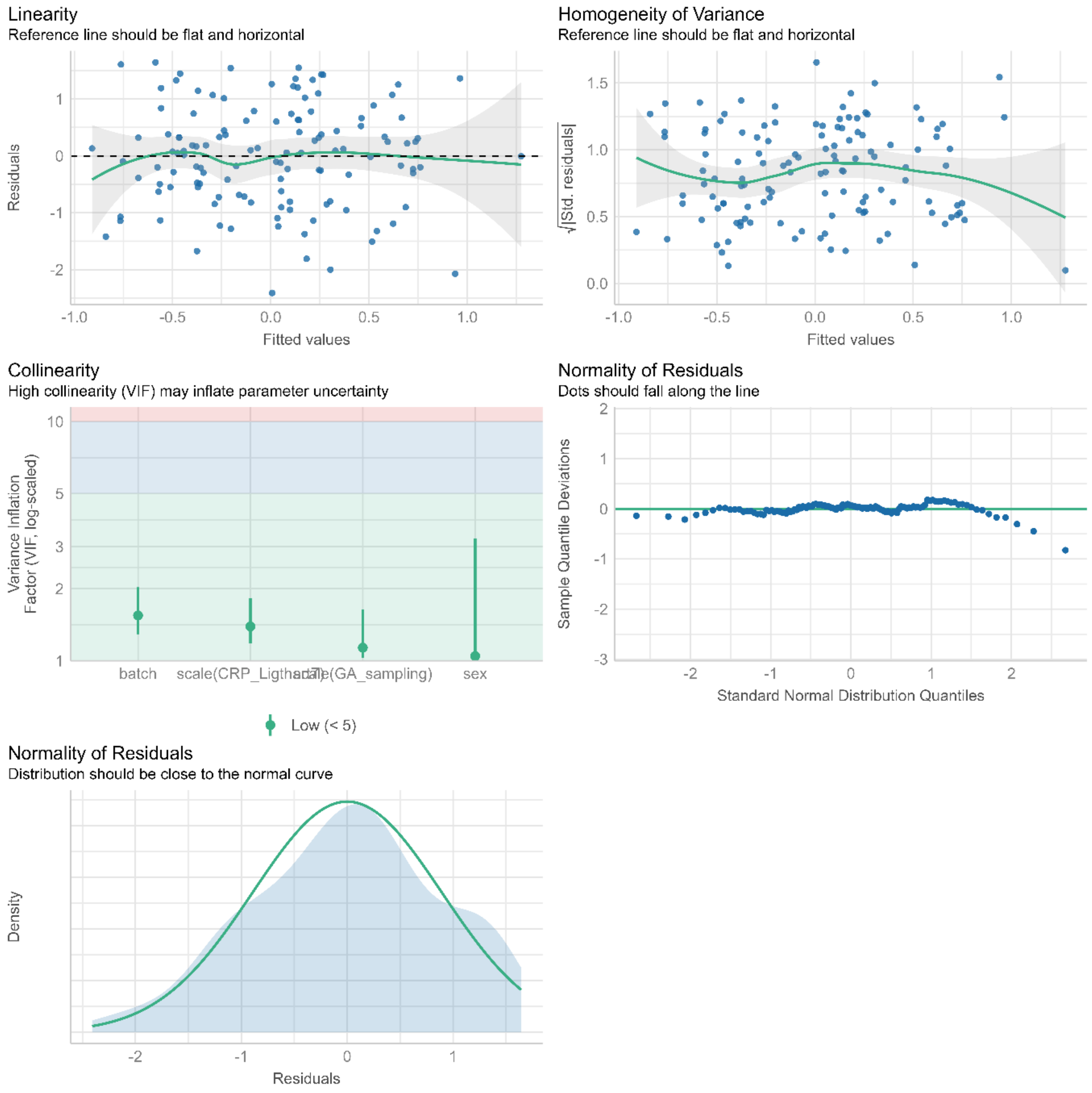


**Supplementary Figure 1**. Model diagnostic metrics for association of DNAm CRP with MSEL Early Learning Composite. Assumptions shown are tested for baseline model = MSEL ELC ~ DNAm CRP + sex + GA sampling + batch

**Supplementary Table 1.** Loadings of the 42 EpiScores onto the first 10 principal components.

| | PC1 | PC2 | PC3 | PC4 | PC5 | PC6 | PC7 | PC8 | PC9 | PC10 |
|---|---|---|---|---|---|---|---|---|---|---|
| Afamin | 0.624 | -0.130 | -0.488 | -0.195 | 0.047 | -0.078 | 0.076 | 0.050 | 0.042 | 0.005 |
| CCL11 | 0.097 | -0.062 | 0.779 | -0.149 | 0.273 | 0.237 | 0.011 | -0.103 | -0.033 | -0.023 |
| CCL18 | 0.011 | 0.274 | -0.201 | 0.311 | -0.028 | -0.248 | 0.423 | -0.305 | 0.215 | 0.244 |
| CCL21 | 0.211 | 0.121 | 0.328 | -0.117 | -0.522 | 0.253 | 0.194 | -0.293 | 0.010 | 0.039 |
| CCL22 | 0.336 | -0.034 | 0.544 | -0.256 | 0.136 | 0.104 | 0.110 | 0.276 | 0.041 | -0.003 |
| CCL25C.C | 0.379 | -0.203 | -0.566 | -0.128 | 0.114 | -0.050 | 0.115 | 0.043 | 0.206 | 0.059 |
| CD163 | 0.440 | 0.715 | -0.081 | -0.052 | 0.101 | 0.223 | -0.079 | -0.024 | -0.001 | 0.033 |
| CD209_antigen | -0.858 | 0.153 | 0.194 | -0.012 | 0.065 | -0.013 | 0.073 | 0.006 | 0.078 | -0.019 |
| CD48_antigen | 0.437 | 0.788 | -0.186 | -0.072 | 0.035 | 0.018 | -0.077 | 0.101 | 0.004 | 0.063 |
| CD6 | 0.323 | 0.038 | 0.254 | -0.482 | 0.120 | 0.269 | 0.117 | -0.055 | -0.025 | -0.143 |
| CD5L | -0.494 | 0.610 | 0.245 | -0.255 | -0.036 | 0.155 | 0.002 | 0.004 | 0.022 | -0.045 |
| ComplementC9 | 0.376 | 0.062 | -0.069 | 0.207 | 0.326 | -0.057 | -0.157 | -0.205 | -0.414 | -0.309 |
| Contactin.4 | -0.460 | -0.213 | -0.534 | 0.194 | 0.289 | 0.253 | 0.182 | 0.062 | -0.116 | 0.157 |
| CXCL10_Olink | -0.131 | 0.693 | 0.109 | 0.219 | 0.021 | -0.078 | -0.004 | -0.087 | 0.046 | 0.160 |
| CXCL9 | 0.060 | 0.737 | -0.110 | 0.416 | -0.007 | -0.129 | -0.033 | 0.126 | 0.004 | -0.014 |
| CRTAM | -0.350 | 0.655 | 0.161 | 0.239 | -0.122 | 0.052 | -0.018 | 0.013 | -0.252 | -0.125 |
| FcRL2 | 0.462 | -0.049 | 0.057 | -0.460 | 0.218 | 0.295 | -0.139 | -0.060 | 0.076 | -0.151 |
| FGF.21 | 0.272 | 0.206 | 0.413 | -0.303 | -0.293 | -0.251 | -0.126 | 0.207 | -0.071 | -0.040 |
| GDF15 | 0.203 | 0.012 | -0.116 | 0.340 | -0.002 | -0.324 | 0.035 | -0.119 | -0.266 | -0.318 |
| GHR | -0.143 | -0.064 | -0.166 | -0.474 | 0.098 | -0.085 | -0.112 | -0.121 | -0.263 | 0.451 |
| HCII | 0.157 | -0.078 | 0.521 | 0.516 | 0.312 | -0.019 | -0.130 | -0.105 | -0.041 | 0.117 |

| | | | | | | | | | | |
|---|---|---|---|---|---|---|---|---|---|---|
| HGF | 0.415 | 0.060 | 0.308 | 0.058 | -0.022 | -0.062 | 0.640 | 0.289 | -0.132 | 0.049 |
| IGFBP.1 | -0.313 | 0.019 | 0.194 | 0.126 | 0.308 | -0.263 | -0.105 | 0.426 | 0.289 | -0.270 |
| IGFBP.4 | 0.515 | 0.021 | 0.275 | -0.345 | -0.174 | -0.149 | -0.267 | -0.098 | 0.108 | -0.116 |
| L.selectin | -0.725 | 0.143 | 0.110 | 0.321 | -0.057 | 0.022 | 0.073 | 0.101 | 0.009 | -0.013 |
| Lactotransferrin | -0.150 | -0.079 | -0.023 | -0.020 | -0.301 | 0.184 | -0.191 | 0.319 | -0.537 | 0.292 |
| FCGR3A | 0.389 | 0.800 | -0.158 | 0.038 | 0.084 | 0.114 | -0.104 | 0.071 | 0.055 | 0.105 |
| MMP.9 | 0.501 | -0.171 | 0.487 | 0.458 | 0.062 | 0.011 | 0.024 | 0.081 | 0.018 | 0.186 |
| NCAM1 | -0.741 | 0.097 | 0.177 | -0.294 | 0.043 | 0.176 | 0.068 | 0.113 | 0.161 | 0.012 |
| PIGR | 0.319 | -0.090 | 0.836 | 0.183 | 0.099 | 0.101 | -0.031 | -0.002 | 0.041 | 0.112 |
| FAP | -0.353 | 0.196 | 0.244 | 0.147 | -0.161 | 0.113 | 0.320 | -0.376 | -0.126 | -0.270 |
| S100.A9 | -0.329 | 0.013 | -0.003 | 0.470 | -0.086 | 0.147 | -0.078 | 0.244 | -0.036 | -0.019 |
| SCGF alpha | 0.347 | -0.181 | -0.333 | 0.517 | -0.236 | 0.477 | -0.054 | 0.058 | 0.159 | -0.073 |
| SCGF beta | 0.449 | -0.185 | -0.372 | 0.411 | -0.271 | 0.443 | -0.076 | 0.100 | 0.187 | -0.087 |
| Semaphorin.3E | -0.609 | 0.133 | -0.411 | -0.395 | -0.149 | -0.012 | 0.075 | -0.023 | 0.099 | -0.119 |
| SKR3 | 0.246 | -0.065 | 0.455 | 0.295 | 0.291 | 0.011 | -0.002 | -0.125 | 0.166 | 0.145 |
| SIGLEC1 | 0.508 | 0.611 | -0.214 | -0.041 | 0.117 | -0.001 | 0.038 | 0.011 | 0.076 | -0.002 |
| SLITRK5 | -0.782 | 0.202 | 0.040 | -0.099 | 0.113 | -0.105 | 0.037 | 0.137 | 0.027 | -0.040 |
| Thrombopoietin receptor | -0.679 | 0.127 | 0.378 | -0.100 | -0.131 | 0.255 | 0.034 | 0.018 | 0.059 | -0.024 |
| Trypsin.2 | 0.269 | 0.087 | 0.389 | 0.031 | -0.596 | -0.333 | -0.185 | -0.022 | 0.217 | 0.122 |
| VCAM1 | 0.189 | 0.802 | -0.215 | -0.127 | 0.181 | 0.109 | -0.027 | -0.032 | 0.015 | 0.036 |
| VEGFA | 0.596 | 0.011 | 0.046 | -0.254 | 0.044 | -0.048 | 0.495 | 0.218 | -0.111 | -0.004 |
| WFIKKN2 | -0.526 | -0.067 | -0.025 | -0.032 | 0.266 | 0.020 | -0.158 | -0.206 | 0.125 | 0.210 |

**Supplementary Table 2**. Demographic and clinical characteristics of participants with neonatal EpiScores and 2-year Bayley Scales of Infant and Toddler Development (Bayley-III) data.

| n | 154 |
|---|---|
| Sex: Female, n (%) | 73 (47.4) |
| Gestational age at birth / weeks, median (range) | 29.21 (22.14, 32.84) |
| Gestational age at DNAm sampling / weeks, median (range) | 40.57 (36.57, 45.86) |
| Neonatal DNAm CRP, median (range) | -0.011 (-0.013, -0.010) |
| Corrected age at Bayley-III assessment / months, median (range) | 24.2 (18.2, 30.2) |
| Bayley-III Cognitive composite, median (range) | 110 (20, 145) |
| Birthweight / g, median (range) | 1150 (370, 2510) |
| Birthweight z-score, median (range) | 0.11 (-3.13, 2.14) |
| Histological chorioamnionitis, n (%) | 43 (30.1) |
| Sepsis, n (%) | 34 (22.1) |
| Bronchopulmonary dysplasia, n (%) | 47 (30.5) |
| Necrotising enterocolitis, n (%) | 3 (1.9) |
| Maternal age / years, median (range) | 32 (17, 44) |
| Maternal BMI, median (range) | 25.30 (16.40, 46.60) |
| Smoked during pregnancy, n (%) | 20 (13.1) |
| Corticosteroid administration in pregnancy, n (%) | 147 (95.5) |
| $MgSO_4$ administration in pregnancy, n (%) | 108 (70.1) |
| Mother university degree, n (%) | 85 (55.2) |
| SIMD rank, median (range) | 4043 (14, 6966) |

SIMD = Scottish Index of Multiple Deprivation.

**Supplementary Table 3.** Association of DNAm CRP with Bayley-III Cognitive Composite in preterm children at 2 years old.

| model | Beta | Lower CI | Upper CI | p | $R^2$ | n |
|---|---|---|---|---|---|---|
| univariable model | 0.014 | -0.122 | 0.150 | 0.862 | -0.007 | 150 |
| baseline model | 0.004 | -0.159 | 0.167 | 0.967 | 0.025 | 150 |
| adjusted model | 0.024 | -0.139 | 0.187 | 0.807 | 0.067 | 150 |

Betas (standardised regression coefficients for DNAm CRP) and 90% confidence intervals are reported. Baseline models are controlled for infant sex, gestational age at sampling and batch. Adjusted models additionally control for gestational age at birth, birthweight Z-score, maternal university education and SIMD.

**Supplementary Table 4.** Association of DNAm CRP with MSEL Early Learning Composite (ELC) in preterm children and term-born children at 5 years old.

| group | model | Beta | Lower CI | Upper CI | p | $R^2$ | n |
|---|---|---|---|---|---|---|---|
| Whole group | univariable model | **-0.273** | -0.417 | -0.129 | **0.002** | 0.067 | 125 |
| | baseline model | **-0.165** | -0.324 | -0.006 | **0.089** | 0.177 | 125 |
| | adjusted model | -0.134 | -0.290 | 0.022 | 0.157 | 0.281 | 125 |
| Preterm | univariable model | **-0.427** | -0.614 | -0.239 | **0.000** | 0.169 | 67 |
| | baseline model | **-0.307** | -0.514 | -0.100 | **0.016** | 0.263 | 67 |
| | adjusted model | **-0.290** | -0.490 | -0.091 | **0.018** | 0.389 | 67 |
| Term | univariable model | 0.055 | -0.168 | 0.279 | 0.680 | -0.015 | 58 |
| | baseline model | **0.296** | 0.035 | 0.556 | **0.063** | 0.184 | 58 |
| | adjusted model | **0.335** | 0.099 | 0.572 | **0.021** | 0.352 | 58 |

Betas (standardised regression coefficients for DNAm CRP) and 90% confidence intervals are reported. Bold text indicate statistically significant associations ($p < 0.10$). Baseline models are controlled for infant sex, gestational age at sampling and batch. Adjusted models additionally control for gestational age at birth, birthweight Z-score, maternal university education and SIMD.

**Supplementary Table 5.** Interaction effects between DNAm CRP and gestational age on MSEL Early Learning Composite in full cohort and preterm only. Standardised betas and p-values are reported from regression models where DNAm CRP is regressed onto MSEL ELC, including an interaction term (DNAm CRP x gestational age at birth (GA birth)), in the model. Bold text denotes p <0.10.

| group | model | Effect | Beta | Lower CI | Upper CI | p | $R^2$ | n |
|---|---|---|---|---|---|---|---|---|
| Whole group | baseline model | DNAm CRP x GA birth | **0.181** | 0.029 | 0.332 | **0.050** | 0.243 | 125 |
| | | DNAm CRP | -0.034 | -0.200 | 0.131 | 0.733 | 0.243 | 125 |
| | adjusted model | DNAm CRP x GA birth | **0.240** | 0.093 | 0.386 | **0.008** | 0.319 | 125 |
| | | DNAm CRP | -0.064 | -0.221 | 0.094 | 0.504 | 0.319 | 125 |
| Preterm | baseline model | DNAm CRP x GA birth | -0.055 | -0.262 | 0.153 | 0.662 | 0.335 | 67 |
| | | DNAm CRP | **-0.226** | -0.430 | -0.022 | **0.069** | 0.335 | 67 |
| | adjusted model | DNAm CRP x GA birth | -0.034 | -0.247 | 0.179 | 0.791 | 0.379 | 67 |
| | | DNAm CRP | **-0.285** | -0.489 | -0.081 | **0.023** | 0.379 | 67 |

Baseline model = MSEL ELC ~ DNAm CRP*GA birth + sex + GA sampling + batch

Adjusted model = MSEL ELC ~ DNAm CRP* GA birth + sex + GA sampling + batch + birthweight Z score + SIMD + maternal university education

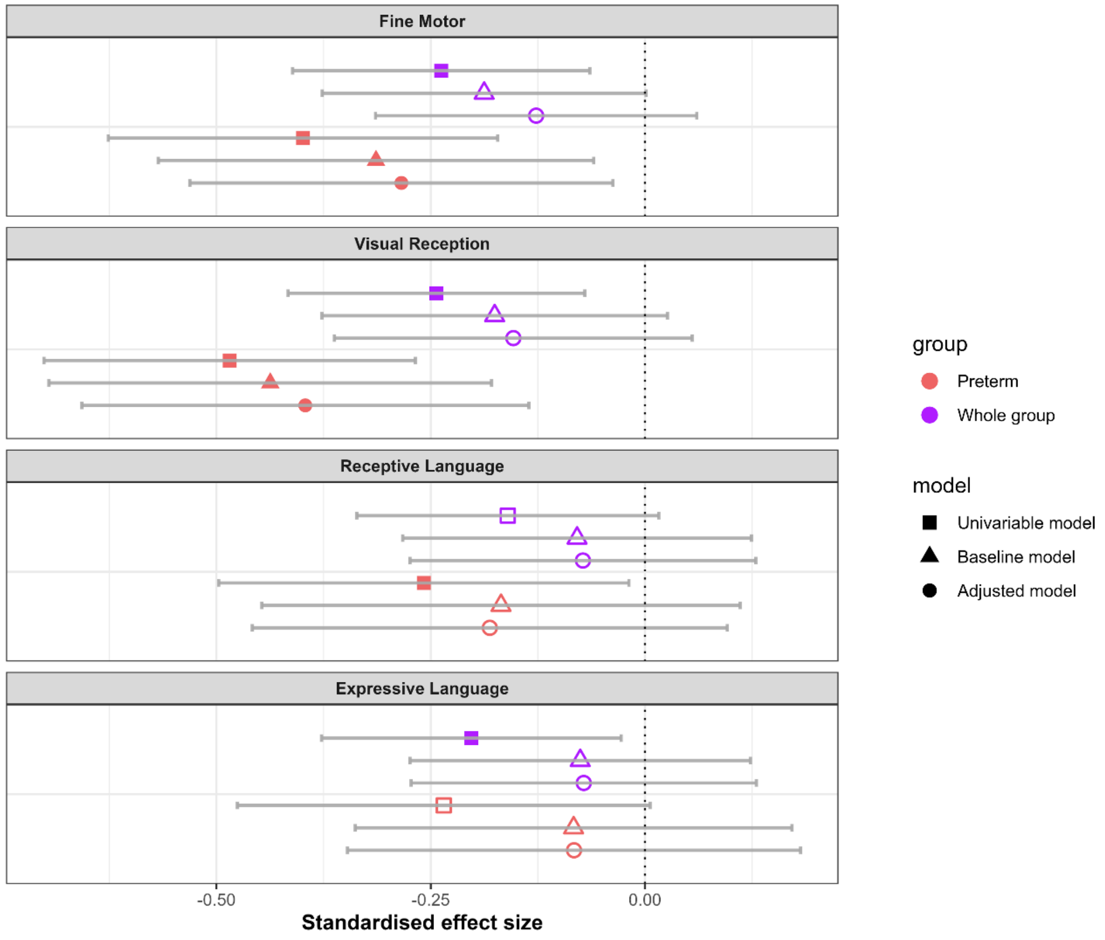


**Supplementary Figure 2.** Associations between DNAm CRP and MSEL subscales in whole group and preterm subgroup. Standardised regression coefficients are shown, where points show standardised coefficients and 95% confidence intervals. Significant associations are denoted by filled shapes corrected for false discovery rate (where q <0.05). Baseline models are controlled for infant sex, gestational age at sampling and batch. Adjusted models additionally control for gestational age at birth, birthweight Z-score, maternal university education and SIMD.

**Supplementary Table 6.** Associations between DNAm CRP and MSEL subscales in exploratory analyses in the full cohort at 5 years old (n = 125). Betas (standardised regression coefficients for DNAm CRP) and 95% confidence intervals are reported. Bold text indicate statistically significant associations which survive FDR correction for multiple comparisons (q <0.05).

| **MSEL subscale** | **model** | **Beta** | **Lower CI** | **Upper CI** | **p** | **$R^2$** | **q** |
|---|---|---|---|---|---|---|---|
| Fine Motor | Univariable model | **-0.238** | -0.411 | -0.064 | 0.008 | 0.049 | **0.015** |
| | Baseline model | -0.188 | -0.377 | 0.001 | 0.052 | 0.188 | 0.175 |
| | Adjusted model | -0.127 | -0.314 | 0.060 | 0.182 | 0.271 | 0.365 |
| Visual Reception | Univariable model | **-0.243** | -0.416 | -0.070 | 0.006 | 0.052 | **0.015** |
| | Baseline model | -0.175 | -0.377 | 0.026 | 0.088 | 0.076 | 0.175 |
| | Adjusted model | -0.154 | -0.362 | 0.055 | 0.148 | 0.096 | 0.365 |
| Receptive Language | Univariable model | -0.160 | -0.336 | 0.016 | 0.074 | 0.018 | 0.074 |
| | Baseline model | -0.079 | -0.283 | 0.124 | 0.442 | 0.059 | 0.453 |
| | Adjusted model | -0.072 | -0.274 | 0.129 | 0.479 | 0.156 | 0.484 |
| Expressive Language | Univariable model | **-0.203** | -0.377 | -0.028 | 0.023 | 0.033 | **0.031** |
| | Baseline model | -0.076 | -0.274 | 0.123 | 0.453 | 0.103 | 0.453 |
| | Adjusted model | -0.071 | -0.273 | 0.130 | 0.484 | 0.159 | 0.484 |

Baseline models are controlled for infant sex, gestational age at sampling and batch. Adjusted models additionally control for gestational age at birth, birthweight Z-score, maternal university education and SIMD.

**Supplementary Table 7.** Associations between DNAm CRP and MSEL subscales in exploratory analyses in the preterm children only at 5 years old (n = 67). Betas (standardised regression coefficients for DNAm CRP) and 95% confidence intervals are reported. Bold text indicate statistically significant associations which survive FDR correction for multiple comparisons (q <0.05).

| MSEL subscale | model | Beta | Lower CI | Upper CI | p | $R^2$ | q |
|---|---|---|---|---|---|---|---|
| Fine Motor | Univariable model | **-0.399** | -0.626 | -0.172 | 0.001 | 0.146 | **0.002** |
| | Baseline model | **-0.314** | -0.568 | -0.060 | 0.016 | 0.226 | **0.033** |
| | Adjusted model | **-0.284** | -0.531 | -0.037 | 0.025 | 0.349 | **0.050** |
| Visual Reception | Univariable model | **-0.485** | -0.701 | -0.268 | 0.000 | 0.223 | **0.000** |
| | Baseline model | **-0.437** | -0.695 | -0.179 | 0.001 | 0.200 | **0.005** |
| | Adjusted model | **-0.396** | -0.657 | -0.136 | 0.004 | 0.274 | **0.014** |
| Receptive Language | Univariable model | **-0.258** | -0.497 | -0.019 | 0.035 | 0.052 | **0.047** |
| | Baseline model | -0.168 | -0.447 | 0.111 | 0.234 | 0.066 | 0.311 |
| | Adjusted model | -0.181 | -0.458 | 0.096 | 0.196 | 0.180 | 0.261 |
| Expressive Language | Univariable model | -0.235 | -0.476 | 0.006 | 0.056 | 0.041 | 0.056 |
| | Baseline model | -0.083 | -0.338 | 0.172 | 0.516 | 0.221 | 0.516 |
| | Adjusted model | -0.083 | -0.347 | 0.182 | 0.533 | 0.254 | 0.533 |

Baseline models are controlled for infant sex, gestational age at sampling and batch. Adjusted models additionally control for gestational age at birth, birthweight Z-score, maternal university education and SIMD.

**Supplementary Table 8.** Williams' test (1959) for differences in magnitudes of standardised DNAm CRP regression coefficients for baseline models between MSEL subscales: fine motor (FM), visual reception (VR), expressive language (EL), receptive language (RL). R1 indicates correlation between subscale 1 and DNAm CRP. R2 indicates correlation between subscale 2 and DNAm CRP. R_common indicates correlation between subscale 1 and subscale 2. Bold text indicate statistically significant differences (t), where $p < 0.05$

| Subscale 1 | Subscale 2 | R1 | R2 | R_common | t | p |
|---|---|---|---|---|---|---|
| FM | VR | -0.314 | -0.437 | 0.526 | 1.118 | 0.264 |
| FM | EL | -0.314 | -0.083 | 0.482 | -1.868 | 0.062 |
| FM | RL | -0.314 | -0.168 | 0.500 | -1.214 | 0.225 |
| VR | EL | -0.437 | -0.083 | 0.546 | **-3.143** | **0.002** |
| VR | RL | -0.437 | -0.168 | 0.609 | **-2.602** | **0.009** |
| EL | RL | -0.083 | -0.168 | 0.566 | 0.735 | 0.462 |

**Supplementary Table 9.** Association of EpiScores with Bayley-III Cognitive Composite in preterm children at 2 years old (n = 150). Betas (standardised regression coefficients for each EpiScore) and 95% confidence intervals are reported. Baseline models are controlled for infant sex, gestational age at sampling and batch. Adjusted models additionally control for gestational age at birth, birthweight Z-score, maternal university education and SIMD.

| EpiScore | model | Beta | Lower CI | Upper CI | p | $R^2$ |
|---|---|---|---|---|---|---|
| **Afamin** | Univariable model | 0.107 | -0.055 | 0.268 | 0.193 | 0.005 |
| | Baseline model | 0.202 | -0.019 | 0.422 | 0.072 | 0.047 |
| | Adjusted model | 0.193 | -0.030 | 0.416 | 0.090 | 0.085 |
| **CCL11** | Univariable model | -0.060 | -0.222 | 0.102 | 0.464 | -0.003 |
| | Baseline model | -0.023 | -0.212 | 0.167 | 0.812 | 0.025 |
| | Adjusted model | -0.012 | -0.199 | 0.175 | 0.900 | 0.066 |
| **CCL18** | Univariable model | 0.180 | 0.020 | 0.340 | 0.028 | 0.026 |
| | Baseline model | 0.198 | 0.036 | 0.361 | 0.017 | 0.063 |
| | Adjusted model | 0.172 | 0.006 | 0.337 | 0.043 | 0.093 |
| **CCL21** | Univariable model | -0.137 | -0.297 | 0.024 | 0.096 | 0.012 |
| | Baseline model | -0.062 | -0.261 | 0.138 | 0.543 | 0.027 |
| | Adjusted model | -0.030 | -0.236 | 0.176 | 0.772 | 0.067 |
| **CCL22** | Univariable model | 0.012 | -0.150 | 0.175 | 0.881 | -0.007 |
| | Baseline model | 0.055 | -0.123 | 0.234 | 0.541 | 0.027 |
| | Adjusted model | 0.029 | -0.148 | 0.206 | 0.746 | 0.067 |
| **CCL25C.C** | Univariable model | 0.007 | -0.155 | 0.170 | 0.928 | -0.007 |
| | Baseline model | -0.048 | -0.255 | 0.160 | 0.651 | 0.026 |
| | Adjusted model | -0.073 | -0.282 | 0.136 | 0.491 | 0.069 |

| | | | | | | |
|---|---|---|---|---|---|---|
| **CD163** | Univariable model | -0.019 | -0.181 | 0.144 | 0.820 | -0.006 |
| | Baseline model | 0.026 | -0.147 | 0.198 | 0.769 | 0.025 |
| | Adjusted model | 0.021 | -0.153 | 0.195 | 0.811 | 0.067 |
| **CD209_antigen** | Univariable model | 0.052 | -0.111 | 0.214 | 0.530 | -0.004 |
| | Baseline model | 0.122 | -0.104 | 0.347 | 0.287 | 0.032 |
| | Adjusted model | 0.085 | -0.141 | 0.312 | 0.458 | 0.070 |
| **CD48_antigen** | Univariable model | 0.106 | -0.056 | 0.267 | 0.197 | 0.005 |
| | Baseline model | 0.135 | -0.039 | 0.309 | 0.128 | 0.040 |
| | Adjusted model | 0.117 | -0.061 | 0.294 | 0.196 | 0.077 |
| **CD6** | Univariable model | 0.006 | -0.156 | 0.169 | 0.938 | -0.007 |
| | Baseline model | 0.002 | -0.163 | 0.167 | 0.980 | 0.025 |
| | Adjusted model | -0.022 | -0.186 | 0.143 | 0.795 | 0.067 |
| **CD5L** | Univariable model | 0.076 | -0.086 | 0.238 | 0.356 | -0.001 |
| | Baseline model | 0.238 | 0.016 | 0.460 | 0.036 | 0.055 |
| | Adjusted model | 0.241 | 0.020 | 0.463 | 0.033 | 0.096 |
| **ComplementC9** | Univariable model | -0.084 | -0.245 | 0.078 | 0.309 | 0.000 |
| | Baseline model | -0.121 | -0.302 | 0.061 | 0.192 | 0.036 |
| | Adjusted model | -0.096 | -0.276 | 0.083 | 0.291 | 0.074 |
| **Contactin.4** | Univariable model | 0.089 | -0.073 | 0.251 | 0.278 | 0.001 |
| | Baseline model | 0.077 | -0.091 | 0.246 | 0.366 | 0.030 |
| | Adjusted model | 0.053 | -0.115 | 0.220 | 0.535 | 0.069 |
| **CXCL10_Olink** | Univariable model | 0.083 | -0.079 | 0.245 | 0.314 | 0.000 |
| | Baseline model | 0.089 | -0.076 | 0.254 | 0.289 | 0.032 |
| | Adjusted model | 0.078 | -0.086 | 0.243 | 0.347 | 0.072 |

| | | | | | | |
|---|---|---|---|---|---|---|
| **CXCL9** | Univariable model | 0.107 | -0.054 | 0.269 | 0.192 | 0.005 |
| | Baseline model | 0.109 | -0.052 | 0.270 | 0.182 | 0.037 |
| | Adjusted model | 0.071 | -0.091 | 0.233 | 0.388 | 0.071 |
| **CRTAM** | Univariable model | -0.050 | -0.212 | 0.112 | 0.545 | -0.004 |
| | Baseline model | -0.019 | -0.200 | 0.163 | 0.839 | 0.025 |
| | Adjusted model | -0.001 | -0.179 | 0.177 | 0.991 | 0.066 |
| **FcRL2** | Univariable model | -0.009 | -0.172 | 0.153 | 0.912 | -0.007 |
| | Baseline model | -0.003 | -0.171 | 0.165 | 0.975 | 0.025 |
| | Adjusted model | -0.021 | -0.190 | 0.147 | 0.803 | 0.067 |
| **FGF.21** | Univariable model | -0.064 | -0.226 | 0.098 | 0.434 | -0.003 |
| | Baseline model | -0.059 | -0.221 | 0.102 | 0.470 | 0.028 |
| | Adjusted model | -0.016 | -0.177 | 0.145 | 0.844 | 0.066 |
| **GDF15** | Univariable model | -0.128 | -0.289 | 0.033 | 0.119 | 0.010 |
| | Baseline model | -0.158 | -0.322 | 0.005 | 0.058 | 0.049 |
| | Adjusted model | -0.151 | -0.314 | 0.012 | 0.069 | 0.088 |
| **GHR** | Univariable model | -0.060 | -0.223 | 0.102 | 0.462 | -0.003 |
| | Baseline model | -0.081 | -0.255 | 0.093 | 0.360 | 0.030 |
| | Adjusted model | -0.098 | -0.269 | 0.074 | 0.262 | 0.075 |
| **HCII** | Univariable model | -0.055 | -0.217 | 0.108 | 0.508 | -0.004 |
| | Baseline model | -0.130 | -0.320 | 0.059 | 0.176 | 0.037 |
| | Adjusted model | -0.126 | -0.317 | 0.066 | 0.197 | 0.077 |
| **HGF** | Univariable model | 0.055 | -0.107 | 0.217 | 0.502 | -0.004 |
| | Baseline model | 0.073 | -0.106 | 0.252 | 0.422 | 0.029 |
| | Adjusted model | 0.053 | -0.127 | 0.232 | 0.563 | 0.068 |

| | | | | | | |
|---|---|---|---|---|---|---|
| **IGFBP.1** | Univariable model | 0.044 | -0.119 | 0.206 | 0.597 | -0.005 |
| | Baseline model | -0.063 | -0.263 | 0.137 | 0.535 | 0.027 |
| | Adjusted model | -0.051 | -0.251 | 0.150 | 0.618 | 0.068 |
| **IGFBP.4** | Univariable model | -0.081 | -0.243 | 0.081 | 0.324 | 0.000 |
| | Baseline model | -0.088 | -0.250 | 0.074 | 0.286 | 0.033 |
| | Adjusted model | -0.072 | -0.233 | 0.088 | 0.375 | 0.072 |
| **L.selectin** | Univariable model | 0.025 | -0.137 | 0.187 | 0.762 | -0.006 |
| | Baseline model | 0.040 | -0.141 | 0.222 | 0.663 | 0.026 |
| | Adjusted model | 0.035 | -0.146 | 0.215 | 0.706 | 0.067 |
| **Lactotransferrin** | Univariable model | 0.042 | -0.121 | 0.204 | 0.613 | -0.005 |
| | Baseline model | 0.061 | -0.105 | 0.227 | 0.467 | 0.028 |
| | Adjusted model | 0.034 | -0.130 | 0.199 | 0.680 | 0.067 |
| **FCGR3A** | Univariable model | 0.126 | -0.036 | 0.287 | 0.126 | 0.009 |
| | Baseline model | 0.156 | -0.012 | 0.324 | 0.069 | 0.047 |
| | Adjusted model | 0.133 | -0.035 | 0.302 | 0.120 | 0.082 |
| **MMP.9** | Univariable model | -0.006 | -0.168 | 0.157 | 0.945 | -0.007 |
| | Baseline model | -0.001 | -0.251 | 0.250 | 0.996 | 0.025 |
| | Adjusted model | -0.031 | -0.279 | 0.218 | 0.808 | 0.067 |
| **NCAM1** | Univariable model | 0.021 | -0.142 | 0.183 | 0.802 | -0.006 |
| | Baseline model | 0.119 | -0.119 | 0.358 | 0.325 | 0.031 |
| | Adjusted model | 0.035 | -0.207 | 0.277 | 0.777 | 0.067 |
| **PIGR** | Univariable model | -0.082 | -0.244 | 0.080 | 0.317 | 0.000 |
| | Baseline model | -0.090 | -0.308 | 0.127 | 0.414 | 0.029 |
| | Adjusted model | -0.090 | -0.305 | 0.125 | 0.407 | 0.071 |

| | | | | | | |
|---|---|---|---|---|---|---|
| **FAP** | Univariable model | -0.014 | -0.177 | 0.148 | 0.863 | -0.007 |
| | Baseline model | 0.040 | -0.137 | 0.216 | 0.656 | 0.026 |
| | Adjusted model | 0.031 | -0.144 | 0.206 | 0.724 | 0.067 |
| **S100.A9** | Univariable model | 0.009 | -0.154 | 0.171 | 0.915 | -0.007 |
| | Baseline model | -0.003 | -0.168 | 0.162 | 0.972 | 0.025 |
| | Adjusted model | -0.028 | -0.195 | 0.138 | 0.739 | 0.067 |
| **SCGF alpha** | Univariable model | 0.098 | -0.064 | 0.260 | 0.233 | 0.003 |
| | Baseline model | 0.157 | -0.038 | 0.352 | 0.114 | 0.042 |
| | Adjusted model | 0.100 | -0.100 | 0.301 | 0.325 | 0.073 |
| **SCGF beta** | Univariable model | 0.070 | -0.092 | 0.233 | 0.391 | -0.002 |
| | Baseline model | 0.138 | -0.067 | 0.342 | 0.185 | 0.037 |
| | Adjusted model | 0.080 | -0.130 | 0.289 | 0.455 | 0.070 |
| **Semaphorin.3E** | Univariable model | 0.012 | -0.150 | 0.175 | 0.880 | -0.007 |
| | Baseline model | 0.015 | -0.208 | 0.238 | 0.893 | 0.025 |
| | Adjusted model | 0.001 | -0.219 | 0.222 | 0.989 | 0.066 |
| **SKR3** | Univariable model | 0.045 | -0.118 | 0.207 | 0.587 | -0.005 |
| | Baseline model | 0.044 | -0.134 | 0.221 | 0.626 | 0.026 |
| | Adjusted model | 0.038 | -0.141 | 0.216 | 0.678 | 0.067 |
| **SIGLEC1** | Univariable model | 0.051 | -0.112 | 0.213 | 0.538 | -0.004 |
| | Baseline model | 0.083 | -0.104 | 0.269 | 0.382 | 0.030 |
| | Adjusted model | 0.060 | -0.130 | 0.250 | 0.531 | 0.069 |
| **SLITRK5** | Univariable model | 0.047 | -0.116 | 0.209 | 0.571 | -0.005 |
| | Baseline model | 0.057 | -0.184 | 0.298 | 0.640 | 0.026 |
| | Adjusted model | 0.041 | -0.197 | 0.279 | 0.732 | 0.067 |

| | | | | | | |
|---|---|---|---|---|---|---|
| **Thrombopoietin_receptor** | Univariable model | -0.071 | -0.233 | 0.092 | 0.391 | -0.002 |
| | Baseline model | -0.046 | -0.284 | 0.192 | 0.702 | 0.026 |
| | Adjusted model | -0.055 | -0.292 | 0.183 | 0.649 | 0.068 |
| **Trypsin.2** | Univariable model | -0.023 | -0.186 | 0.139 | 0.775 | -0.006 |
| | Baseline model | -0.018 | -0.183 | 0.146 | 0.826 | 0.025 |
| | Adjusted model | 0.013 | -0.150 | 0.176 | 0.875 | 0.066 |
| **VCAM1** | Univariable model | 0.090 | -0.072 | 0.252 | 0.274 | 0.001 |
| | Baseline model | 0.147 | -0.024 | 0.318 | 0.091 | 0.044 |
| | Adjusted model | 0.144 | -0.029 | 0.317 | 0.101 | 0.084 |
| **VEGFA** | Univariable model | 0.042 | -0.120 | 0.204 | 0.611 | -0.005 |
| | Baseline model | 0.052 | -0.134 | 0.239 | 0.580 | 0.027 |
| | Adjusted model | 0.033 | -0.153 | 0.220 | 0.726 | 0.067 |
| **WFIKKN2** | Univariable model | 0.104 | -0.057 | 0.266 | 0.205 | 0.004 |
| | Baseline model | 0.097 | -0.091 | 0.284 | 0.310 | 0.032 |
| | Adjusted model | 0.051 | -0.138 | 0.239 | 0.594 | 0.068 |

**Supplementary Table 10.** Association of EpiScores with MSEL Early Learning Composite (ELC) in the full cohort at 5 years old (n = 125). Betas (standardised regression coefficients for each EpiScore) and 95% confidence intervals are reported. Bold text indicate statistically significant associations (Bonferroni-adjusted significance threshold p <0.01). Baseline models are controlled for infant sex, gestational age at sampling and batch. Adjusted models additionally control for gestational age at birth, birthweight Z-score, maternal university education and SIMD.

| EpiScore | model | Beta | Lower CI | Upper CI | p | $R^2$ |
|---|---|---|---|---|---|---|
| **Afamin** | Univariable model | -0.180 | -0.355 | -0.004 | 0.045 | 0.024 |
| | Baseline model | 0.028 | -0.178 | 0.234 | 0.788 | 0.158 |
| | Adjusted model | 0.035 | -0.161 | 0.231 | 0.724 | 0.270 |
| **CCL11** | Univariable model | 0.108 | -0.069 | 0.285 | 0.231 | 0.004 |
| | Baseline model | -0.078 | -0.272 | 0.116 | 0.428 | 0.161 |
| | Adjusted model | -0.063 | -0.245 | 0.119 | 0.497 | 0.272 |
| **CCL18** | Univariable model | 0.099 | -0.078 | 0.277 | 0.270 | 0.002 |
| | Baseline model | 0.088 | -0.076 | 0.253 | 0.289 | 0.165 |
| | Adjusted model | 0.100 | -0.054 | 0.255 | 0.202 | 0.279 |
| **CCL21** | Univariable model | -0.211 | -0.386 | -0.037 | 0.018 | 0.037 |
| | Baseline model | -0.151 | -0.349 | 0.048 | 0.136 | 0.173 |
| | Adjusted model | -0.097 | -0.290 | 0.095 | 0.319 | 0.275 |
| **CCL22** | Univariable model | -0.009 | -0.187 | 0.170 | 0.922 | -0.008 |
| | Baseline model | -0.097 | -0.263 | 0.069 | 0.248 | 0.166 |
| | Adjusted model | -0.087 | -0.247 | 0.072 | 0.280 | 0.276 |
| **CCL25C.C** | Univariable model | -0.135 | -0.312 | 0.042 | 0.133 | 0.010 |

| | | | | | | |
|---|---|---|---|---|---|---|
| | Baseline model | 0.129 | -0.082 | 0.341 | 0.228 | 0.167 |
| | Adjusted model | -0.017 | -0.237 | 0.203 | 0.877 | 0.269 |
| **CD163** | Univariable model | -0.077 | -0.255 | 0.101 | 0.395 | -0.002 |
| | Baseline model | -0.074 | -0.238 | 0.090 | 0.372 | 0.163 |
| | Adjusted model | -0.020 | -0.187 | 0.147 | 0.812 | 0.269 |
| **CD209_antigen** | Univariable model | **0.267** | 0.095 | 0.439 | **0.003** | 0.064 |
| | Baseline model | 0.079 | -0.122 | 0.281 | 0.438 | 0.161 |
| | Adjusted model | 0.002 | -0.196 | 0.200 | 0.985 | 0.269 |
| **CD48_antigen** | Univariable model | -0.170 | -0.346 | 0.006 | 0.058 | 0.021 |
| | Baseline model | -0.143 | -0.314 | 0.028 | 0.100 | 0.176 |
| | Adjusted model | -0.111 | -0.285 | 0.062 | 0.207 | 0.279 |
| **CD6** | Univariable model | -0.031 | -0.209 | 0.148 | 0.734 | -0.007 |
| | Baseline model | -0.035 | -0.199 | 0.129 | 0.671 | 0.158 |
| | Adjusted model | -0.018 | -0.176 | 0.141 | 0.826 | 0.269 |
| **CD5L** | Univariable model | 0.027 | -0.152 | 0.205 | 0.767 | -0.007 |
| | Baseline model | -0.198 | -0.395 | 0.000 | 0.050 | 0.184 |
| | Adjusted model | -0.166 | -0.361 | 0.030 | 0.096 | 0.286 |
| **ComplementC9** | Univariable model | -0.096 | -0.274 | 0.081 | 0.285 | 0.001 |
| | Baseline model | -0.046 | -0.220 | 0.127 | 0.596 | 0.159 |
| | Adjusted model | -0.045 | -0.218 | 0.128 | 0.609 | 0.270 |
| **Contactin.4** | Univariable model | 0.105 | -0.072 | 0.283 | 0.242 | 0.003 |
| | Baseline model | 0.157 | -0.005 | 0.319 | 0.057 | 0.182 |
| | Adjusted model | 0.091 | -0.075 | 0.257 | 0.281 | 0.276 |
| **CXCL10_Olink** | Univariable model | 0.017 | -0.162 | 0.195 | 0.855 | -0.008 |

| | | | | | | |
|---|---|---|---|---|---|---|
| | Baseline model | -0.072 | -0.240 | 0.095 | 0.394 | 0.162 |
| | Adjusted model | -0.001 | -0.165 | 0.163 | 0.994 | 0.269 |
| **CXCL9** | Univariable model | 0.056 | -0.122 | 0.234 | 0.535 | -0.005 |
| | Baseline model | -0.003 | -0.173 | 0.166 | 0.969 | 0.157 |
| | Adjusted model | 0.086 | -0.077 | 0.250 | 0.298 | 0.276 |
| **CRTAM** | Univariable model | -0.102 | -0.279 | 0.076 | 0.258 | 0.002 |
| | Baseline model | **-0.219** | -0.384 | -0.054 | **0.010** | 0.203 |
| | Adjusted model | -0.123 | -0.290 | 0.045 | 0.149 | 0.282 |
| **FcRL2** | Univariable model | -0.065 | -0.243 | 0.113 | 0.472 | -0.004 |
| | Baseline model | -0.023 | -0.189 | 0.142 | 0.779 | 0.158 |
| | Adjusted model | -0.037 | -0.195 | 0.121 | 0.643 | 0.270 |
| **FGF.21** | Univariable model | -0.041 | -0.220 | 0.137 | 0.648 | -0.006 |
| | Baseline model | -0.101 | -0.266 | 0.064 | 0.229 | 0.167 |
| | Adjusted model | -0.057 | -0.217 | 0.103 | 0.483 | 0.272 |
| **GDF15** | Univariable model | -0.155 | -0.331 | 0.021 | 0.084 | 0.016 |
| | Baseline model | -0.099 | -0.267 | 0.068 | 0.244 | 0.167 |
| | Adjusted model | -0.057 | -0.217 | 0.104 | 0.485 | 0.272 |
| **GHR** | Univariable model | 0.029 | -0.149 | 0.208 | 0.747 | -0.007 |
| | Baseline model | 0.053 | -0.110 | 0.216 | 0.522 | 0.160 |
| | Adjusted model | 0.005 | -0.151 | 0.162 | 0.946 | 0.269 |
| **HCII** | Univariable model | 0.060 | -0.118 | 0.238 | 0.509 | -0.005 |
| | Baseline model | -0.245 | -0.443 | -0.048 | 0.015 | 0.197 |
| | Adjusted model | -0.186 | -0.377 | 0.004 | 0.055 | 0.292 |
| **HGF** | Univariable model | -0.228 | -0.401 | -0.054 | 0.011 | 0.044 |

| | | | | | | |
|---|---|---|---|---|---|---|
| | Baseline model | -0.159 | -0.325 | 0.006 | 0.059 | 0.182 |
| | Adjusted model | -0.086 | -0.248 | 0.076 | 0.296 | 0.276 |
| **IGFBP.1** | Univariable model | 0.166 | -0.010 | 0.342 | 0.064 | 0.020 |
| | Baseline model | -0.003 | -0.187 | 0.180 | 0.972 | 0.157 |
| | Adjusted model | -0.014 | -0.186 | 0.158 | 0.870 | 0.269 |
| **IGFBP.4** | Univariable model | -0.091 | -0.269 | 0.087 | 0.313 | 0.000 |
| | Baseline model | -0.084 | -0.247 | 0.079 | 0.310 | 0.164 |
| | Adjusted model | -0.062 | -0.221 | 0.098 | 0.444 | 0.273 |
| **L.selectin** | Univariable model | 0.166 | -0.010 | 0.342 | 0.064 | 0.020 |
| | Baseline model | 0.075 | -0.096 | 0.247 | 0.386 | 0.162 |
| | Adjusted model | 0.079 | -0.084 | 0.243 | 0.339 | 0.275 |
| **Lactotransferrin** | Univariable model | 0.119 | -0.058 | 0.296 | 0.187 | 0.006 |
| | Baseline model | 0.160 | -0.006 | 0.326 | 0.059 | 0.182 |
| | Adjusted model | 0.109 | -0.056 | 0.274 | 0.192 | 0.280 |
| **FCGR3A** | Univariable model | -0.111 | -0.288 | 0.067 | 0.219 | 0.004 |
| | Baseline model | -0.098 | -0.267 | 0.070 | 0.249 | 0.166 |
| | Adjusted model | -0.047 | -0.214 | 0.120 | 0.579 | 0.271 |
| **MMP.9** | Univariable model | 0.046 | -0.132 | 0.225 | 0.607 | -0.006 |
| | Baseline model | -0.032 | -0.202 | 0.139 | 0.715 | 0.158 |
| | Adjusted model | 0.009 | -0.151 | 0.169 | 0.913 | 0.269 |
| **NCAM1** | Univariable model | 0.228 | 0.054 | 0.402 | 0.011 | 0.044 |
| | Baseline model | 0.094 | -0.115 | 0.303 | 0.374 | 0.163 |
| | Adjusted model | -0.011 | -0.218 | 0.196 | 0.916 | 0.269 |
| **PIGR** | Univariable model | 0.125 | -0.052 | 0.302 | 0.165 | 0.008 |

| | | | | | | |
|---|---|---|---|---|---|---|
| | Baseline model | -0.209 | -0.425 | 0.007 | 0.058 | 0.182 |
| | Adjusted model | -0.115 | -0.324 | 0.094 | 0.279 | 0.276 |
| **FAP** | Univariable model | 0.000 | -0.179 | 0.178 | 0.996 | -0.008 |
| | Baseline model | -0.048 | -0.235 | 0.139 | 0.615 | 0.159 |
| | Adjusted model | -0.007 | -0.183 | 0.168 | 0.933 | 0.269 |
| **S100.A9** | Univariable model | 0.112 | -0.065 | 0.290 | 0.213 | 0.005 |
| | Baseline model | 0.056 | -0.111 | 0.222 | 0.510 | 0.160 |
| | Adjusted model | 0.045 | -0.115 | 0.205 | 0.576 | 0.271 |
| **SCGF alpha** | Univariable model | -0.047 | -0.225 | 0.131 | 0.602 | -0.006 |
| | Baseline model | 0.197 | 0.015 | 0.379 | 0.034 | 0.188 |
| | Adjusted model | 0.163 | -0.009 | 0.335 | 0.063 | 0.290 |
| **SCGF beta** | Univariable model | -0.106 | -0.284 | 0.071 | 0.239 | 0.003 |
| | Baseline model | 0.196 | -0.003 | 0.395 | 0.054 | 0.183 |
| | Adjusted model | 0.171 | -0.016 | 0.358 | 0.072 | 0.289 |
| **Semaphorin.3E** | Univariable model | 0.085 | -0.093 | 0.263 | 0.346 | -0.001 |
| | Baseline model | 0.125 | -0.039 | 0.288 | 0.133 | 0.173 |
| | Adjusted model | 0.063 | -0.094 | 0.220 | 0.427 | 0.273 |
| **SKR3** | Univariable model | 0.035 | -0.143 | 0.214 | 0.697 | -0.007 |
| | Baseline model | -0.151 | -0.331 | 0.029 | 0.100 | 0.176 |
| | Adjusted model | -0.081 | -0.257 | 0.096 | 0.366 | 0.274 |
| **SIGLEC1** | Univariable model | -0.196 | -0.371 | -0.021 | 0.029 | 0.030 |
| | Baseline model | -0.145 | -0.317 | 0.028 | 0.099 | 0.176 |
| | Adjusted model | -0.081 | -0.262 | 0.100 | 0.377 | 0.274 |
| **SLITRK5** | Univariable model | **0.242** | 0.068 | 0.415**0.** | **0.007** | 0.051 |

| | | | | | | |
|---|---|---|---|---|---|---|
| | Baseline model | 0.064 | -0.122 | 0.251 | 0.497 | 0.160 |
| | Adjusted model | 0.058 | -0.123 | 0.239 | 0.528 | 0.271 |
| **Thrombopoietin_receptor** | Univariable model | 0.205 | 0.030 | 0.379 | 0.022 | 0.034 |
| | Baseline model | 0.030 | -0.205 | 0.264 | 0.803 | 0.157 |
| | Adjusted model | -0.063 | -0.295 | 0.168 | 0.589 | 0.271 |
| **Trypsin.2** | Univariable model | -0.082 | -0.260 | 0.096 | 0.362 | -0.001 |
| | Baseline model | -0.097 | -0.261 | 0.066 | 0.242 | 0.167 |
| | Adjusted model | -0.012 | -0.170 | 0.147 | 0.883 | 0.269 |
| **VCAM1** | Univariable model | -0.115 | -0.292 | 0.063 | 0.203 | 0.005 |
| | Baseline model | -0.094 | -0.261 | 0.072 | 0.264 | 0.166 |
| | Adjusted model | -0.049 | -0.218 | 0.120 | 0.566 | 0.271 |
| **VEGFA** | Univariable model | -0.221 | -0.395 | -0.047 | 0.013 | 0.041 |
| | Baseline model | -0.104 | -0.277 | 0.069 | 0.238 | 0.167 |
| | Adjusted model | -0.052 | -0.218 | 0.114 | 0.536 | 0.271 |
| **WFIKKN2** | Univariable model | 0.192 | 0.017 | 0.367 | 0.032 | 0.029 |
| | Baseline model | 0.108 | -0.058 | 0.274 | 0.199 | 0.169 |
| | Adjusted model | 0.042 | -0.118 | 0.202 | 0.606 | 0.271 |

**Supplementary Table 11.** Associations between EpiScores and MSEL Early Learning Composite, adjusting for number of inflammatory exposures in preterm children only. Betas (standardised regression coefficients for each EpiScore) and 95% confidence intervals are reported. Bold text indicate statistically significant associations (Bonferroni-adjusted significance threshold p <0.01). Baseline models are controlled for infant sex, gestational age at sampling and batch. Adjusted models additionally control for gestational age at birth, birthweight Z-score, maternal university education and SIMD.

| EpiScore | model | Beta | Lower CI | Upper CI | p | $R^2$ | n |
|---|---|---|---|---|---|---|---|
| **Afamin** | baseline model | 0.209 | -0.078 | 0.495 | 0.151 | 0.217 | 67 |
| | baseline model + inflammatory exposures | 0.205 | -0.088 | 0.498 | 0.167 | 0.219 | 63 |
| | adjusted model + inflammatory exposures | 0.117 | -0.159 | 0.393 | 0.398 | 0.334 | 63 |
| **CCL11** | baseline model | 0.241 | -0.028 | 0.509 | 0.078 | 0.230 | 67 |
| | baseline model + inflammatory exposures | 0.232 | -0.043 | 0.506 | 0.096 | 0.231 | 63 |
| | adjusted model + inflammatory exposures | 0.239 | -0.012 | 0.489 | 0.062 | 0.368 | 63 |
| **CCL18** | baseline model | 0.093 | -0.129 | 0.316 | 0.406 | 0.199 | 67 |
| | baseline model + inflammatory exposures | 0.129 | -0.108 | 0.367 | 0.281 | 0.208 | 63 |
| | adjusted model + inflammatory exposures | 0.163 | -0.055 | 0.381 | 0.140 | 0.352 | 63 |
| **CCL21** | baseline model | -0.200 | -0.486 | 0.087 | 0.169 | 0.215 | 67 |
| | baseline model + inflammatory exposures | -0.233 | -0.539 | 0.074 | 0.134 | 0.223 | 63 |
| | adjusted model + inflammatory exposures | -0.197 | -0.479 | 0.085 | 0.168 | 0.349 | 63 |
| **CCL22** | baseline model | 0.028 | -0.197 | 0.252 | 0.807 | 0.191 | 67 |
| | baseline model + inflammatory exposures | 0.029 | -0.207 | 0.264 | 0.809 | 0.193 | 63 |
| | adjusted model + inflammatory exposures | 0.004 | -0.213 | 0.222 | 0.968 | 0.325 | 63 |
| **CCL25C.C** | baseline model | 0.170 | -0.130 | 0.470 | 0.262 | 0.207 | 67 |
| | baseline model + inflammatory exposures | 0.158 | -0.157 | 0.473 | 0.320 | 0.206 | 63 |
| | adjusted model + inflammatory exposures | 0.025 | -0.275 | 0.325 | 0.868 | 0.325 | 63 |
| **CD163** | baseline model | 0.002 | -0.231 | 0.235 | 0.986 | 0.190 | 67 |
| | baseline model + inflammatory exposures | -0.041 | -0.290 | 0.207 | 0.742 | 0.194 | 63 |
| | adjusted model + inflammatory exposures | -0.012 | -0.251 | 0.226 | 0.918 | 0.325 | 63 |
| **CD209_antigen** | baseline model | 0.096 | -0.180 | 0.373 | 0.489 | 0.197 | 67 |
| | baseline model + inflammatory exposures | 0.130 | -0.156 | 0.415 | 0.366 | 0.204 | 63 |
| | adjusted model + inflammatory exposures | 0.130 | -0.136 | 0.396 | 0.330 | 0.337 | 63 |
| **CD48_antigen** | baseline model | -0.075 | -0.328 | 0.178 | 0.558 | 0.195 | 67 |
| | baseline model + inflammatory exposures | -0.109 | -0.376 | 0.158 | 0.418 | 0.201 | 63 |
| | adjusted model + inflammatory exposures | -0.133 | -0.388 | 0.123 | 0.303 | 0.339 | 63 |
| **CD6** | baseline model | 0.186 | -0.042 | 0.413 | 0.108 | 0.224 | 67 |

| | | | | | | | |
|---|---|---|---|---|---|---|---|
| | baseline model + inflammatory exposures | 0.192 | -0.044 | 0.428 | 0.109 | 0.228 | 63 |
| | adjusted model + inflammatory exposures | 0.103 | -0.124 | 0.330 | 0.365 | 0.335 | 63 |
| **CD5L** | baseline model | -0.002 | -0.259 | 0.255 | 0.988 | 0.190 | 67 |
| | baseline model + inflammatory exposures | -0.032 | -0.297 | 0.233 | 0.810 | 0.193 | 63 |
| | adjusted model + inflammatory exposures | 0.028 | -0.228 | 0.285 | 0.825 | 0.326 | 63 |
| **ComplementC9** | baseline model | -0.152 | -0.396 | 0.092 | 0.217 | 0.210 | 67 |
| | baseline model + inflammatory exposures | -0.139 | -0.396 | 0.118 | 0.283 | 0.208 | 63 |
| | adjusted model + inflammatory exposures | -0.124 | -0.386 | 0.138 | 0.348 | 0.336 | 63 |
| **Contactin.4** | baseline model | 0.104 | -0.125 | 0.333 | 0.369 | 0.201 | 67 |
| | baseline model + inflammatory exposures | 0.122 | -0.121 | 0.365 | 0.318 | 0.206 | 63 |
| | adjusted model + inflammatory exposures | 0.131 | -0.096 | 0.359 | 0.252 | 0.342 | 63 |
| **CXCL10_Olink** | baseline model | -0.097 | -0.326 | 0.131 | 0.397 | 0.200 | 67 |
| | baseline model + inflammatory exposures | -0.130 | -0.368 | 0.108 | 0.278 | 0.209 | 63 |
| | adjusted model + inflammatory exposures | -0.013 | -0.249 | 0.224 | 0.915 | 0.325 | 63 |
| **CXCL9** | baseline model | -0.105 | -0.336 | 0.125 | 0.365 | 0.201 | 67 |
| | baseline model + inflammatory exposures | -0.110 | -0.349 | 0.129 | 0.360 | 0.204 | 63 |
| | adjusted model + inflammatory exposures | -0.041 | -0.266 | 0.184 | 0.717 | 0.327 | 63 |
| **CRTAM** | baseline model | **-0.307** | -0.523 | -0.090 | **0.006** | 0.283 | 67 |
| | baseline model + inflammatory exposures | **-0.307** | -0.531 | -0.083 | **0.008** | 0.286 | 63 |
| | adjusted model + inflammatory exposures | -0.265 | -0.477 | -0.052 | 0.016 | 0.396 | 63 |
| **FcRL2** | baseline model | 0.195 | -0.030 | 0.420 | 0.088 | 0.228 | 67 |
| | baseline model + inflammatory exposures | 0.180 | -0.053 | 0.413 | 0.127 | 0.225 | 63 |
| | adjusted model + inflammatory exposures | 0.103 | -0.123 | 0.329 | 0.364 | 0.336 | 63 |
| **FGF.21** | baseline model | -0.087 | -0.314 | 0.141 | 0.450 | 0.198 | 67 |
| | baseline model + inflammatory exposures | -0.106 | -0.341 | 0.130 | 0.372 | 0.203 | 63 |
| | adjusted model + inflammatory exposures | -0.076 | -0.302 | 0.150 | 0.502 | 0.331 | 63 |
| **GDF15** | baseline model | -0.218 | -0.438 | 0.001 | 0.051 | 0.239 | 67 |
| | baseline model + inflammatory exposures | -0.211 | -0.439 | 0.016 | 0.068 | 0.238 | 63 |
| | adjusted model + inflammatory exposures | -0.163 | -0.378 | 0.051 | 0.133 | 0.353 | 63 |

| | | | | | | | |
|---|---|---|---|---|---|---|---|
| **GHR** | baseline model | 0.168 | -0.053 | 0.389 | 0.133 | 0.220 | 67 |
| | baseline model + inflammatory exposures | 0.137 | -0.101 | 0.375 | 0.253 | 0.210 | 63 |
| | adjusted model + inflammatory exposures | 0.023 | -0.214 | 0.260 | 0.844 | 0.326 | 63 |
| **HCII** | baseline model | -0.273 | -0.544 | -0.002 | 0.048 | 0.240 | 67 |
| | baseline model + inflammatory exposures | -0.265 | -0.546 | 0.016 | 0.064 | 0.239 | 63 |
| | adjusted model + inflammatory exposures | -0.135 | -0.416 | 0.145 | 0.339 | 0.337 | 63 |
| **HGF** | baseline model | -0.102 | -0.328 | 0.124 | 0.373 | 0.201 | 67 |
| | baseline model + inflammatory exposures | -0.128 | -0.360 | 0.104 | 0.273 | 0.209 | 63 |
| | adjusted model + inflammatory exposures | -0.078 | -0.298 | 0.142 | 0.481 | 0.331 | 63 |
| **IGFBP.1** | baseline model | 0.003 | -0.249 | 0.255 | 0.979 | 0.190 | 67 |
| | baseline model + inflammatory exposures | 0.018 | -0.248 | 0.285 | 0.891 | 0.192 | 63 |
| | adjusted model + inflammatory exposures | 0.060 | -0.184 | 0.305 | 0.622 | 0.328 | 63 |
| **IGFBP.4** | baseline model | -0.024 | -0.252 | 0.205 | 0.837 | 0.191 | 67 |
| | baseline model + inflammatory exposures | -0.016 | -0.249 | 0.218 | 0.894 | 0.192 | 63 |
| | adjusted model + inflammatory exposures | -0.083 | -0.304 | 0.138 | 0.452 | 0.332 | 63 |
| **L.selectin** | baseline model | -0.035 | -0.280 | 0.209 | 0.773 | 0.191 | 67 |
| | baseline model + inflammatory exposures | -0.018 | -0.270 | 0.233 | 0.884 | 0.192 | 63 |
| | adjusted model + inflammatory exposures | 0.038 | -0.199 | 0.274 | 0.752 | 0.326 | 63 |
| **Lactotransferrin** | baseline model | -0.011 | -0.258 | 0.237 | 0.931 | 0.190 | 67 |
| | baseline model + inflammatory exposures | 0.002 | -0.259 | 0.262 | 0.990 | 0.192 | 63 |
| | adjusted model + inflammatory exposures | -0.041 | -0.282 | 0.199 | 0.732 | 0.327 | 63 |
| **FCGR3A** | baseline model | -0.068 | -0.304 | 0.168 | 0.565 | 0.195 | 67 |
| | baseline model + inflammatory exposures | -0.089 | -0.334 | 0.157 | 0.474 | 0.199 | 63 |
| | adjusted model + inflammatory exposures | -0.082 | -0.316 | 0.152 | 0.486 | 0.331 | 63 |
| **MMP.9** | baseline model | -0.146 | -0.377 | 0.085 | 0.210 | 0.211 | 67 |
| | baseline model + inflammatory exposures | -0.138 | -0.376 | 0.101 | 0.253 | 0.210 | 63 |
| | adjusted model + inflammatory exposures | -0.102 | -0.326 | 0.123 | 0.368 | 0.335 | 63 |
| **NCAM1** | baseline model | 0.294 | 0.013 | 0.574 | 0.040 | 0.244 | 67 |
| | baseline model + inflammatory exposures | 0.274 | -0.020 | 0.568 | 0.067 | 0.238 | 63 |

| | | | | | | | |
|---|---|---|---|---|---|---|---|
| | adjusted model + inflammatory exposures | 0.303 | 0.027 | 0.578 | 0.032 | 0.382 | 63 |
| **PIGR** | baseline model | -0.130 | -0.443 | 0.182 | 0.408 | 0.199 | 67 |
| | baseline model + inflammatory exposures | -0.135 | -0.456 | 0.187 | 0.405 | 0.202 | 63 |
| | adjusted model + inflammatory exposures | -0.104 | -0.402 | 0.195 | 0.488 | 0.331 | 63 |
| **FAP** | baseline model | -0.012 | -0.253 | 0.229 | 0.921 | 0.190 | 67 |
| | baseline model + inflammatory exposures | 0.070 | -0.191 | 0.331 | 0.595 | 0.196 | 63 |
| | adjusted model + inflammatory exposures | 0.088 | -0.153 | 0.329 | 0.469 | 0.332 | 63 |
| **S100.A9** | baseline model | -0.217 | -0.448 | 0.014 | 0.065 | 0.234 | 67 |
| | baseline model + inflammatory exposures | -0.210 | -0.447 | 0.027 | 0.081 | 0.234 | 63 |
| | adjusted model + inflammatory exposures | -0.124 | -0.358 | 0.110 | 0.293 | 0.339 | 63 |
| **SCGF alpha** | baseline model | 0.115 | -0.143 | 0.373 | 0.376 | 0.201 | 67 |
| | baseline model + inflammatory exposures | 0.117 | -0.160 | 0.395 | 0.401 | 0.202 | 63 |
| | adjusted model + inflammatory exposures | 0.046 | -0.226 | 0.318 | 0.737 | 0.326 | 63 |
| **SCGF beta** | baseline model | 0.165 | -0.120 | 0.450 | 0.251 | 0.207 | 67 |
| | baseline model + inflammatory exposures | 0.163 | -0.145 | 0.472 | 0.294 | 0.208 | 63 |
| | adjusted model + inflammatory exposures | 0.093 | -0.202 | 0.388 | 0.531 | 0.330 | 63 |
| **Semaphorin.3E** | baseline model | 0.134 | -0.086 | 0.355 | 0.228 | 0.209 | 67 |
| | baseline model + inflammatory exposures | 0.138 | -0.089 | 0.366 | 0.229 | 0.212 | 63 |
| | adjusted model + inflammatory exposures | 0.088 | -0.128 | 0.305 | 0.417 | 0.333 | 63 |
| **SKR3** | baseline model | -0.077 | -0.333 | 0.179 | 0.550 | 0.195 | 67 |
| | baseline model + inflammatory exposures | -0.099 | -0.366 | 0.168 | 0.462 | 0.200 | 63 |
| | adjusted model + inflammatory exposures | -0.022 | -0.277 | 0.233 | 0.865 | 0.325 | 63 |
| **SIGLEC1** | baseline model | -0.113 | -0.397 | 0.170 | 0.427 | 0.199 | 67 |
| | baseline model + inflammatory exposures | -0.131 | -0.421 | 0.159 | 0.370 | 0.203 | 63 |
| | adjusted model + inflammatory exposures | -0.037 | -0.323 | 0.250 | 0.799 | 0.326 | 63 |
| **SLITRK5** | baseline model | 0.000 | -0.255 | 0.255 | 1.000 | 0.190 | 67 |
| | baseline model + inflammatory exposures | 0.044 | -0.223 | 0.312 | 0.742 | 0.194 | 63 |
| | adjusted model + inflammatory exposures | 0.067 | -0.188 | 0.322 | 0.599 | 0.329 | 63 |
| | baseline model | 0.118 | -0.204 | 0.441 | 0.466 | 0.197 | 67 |

| | | | | | | | |
|---|---|---|---|---|---|---|---|
| **Thrombopoietin receptor** | baseline model + inflammatory exposures | 0.076 | -0.266 | 0.418 | 0.658 | 0.195 | 63 |
| | adjusted model + inflammatory exposures | 0.072 | -0.257 | 0.400 | 0.663 | 0.327 | 63 |
| **Trypsin.2** | baseline model | -0.165 | -0.392 | 0.063 | 0.153 | 0.217 | 67 |
| | baseline model + inflammatory exposures | -0.159 | -0.396 | 0.078 | 0.184 | 0.217 | 63 |
| | adjusted model + inflammatory exposures | -0.170 | -0.405 | 0.066 | 0.154 | 0.351 | 63 |
| **VCAM1** | baseline model | 0.048 | -0.192 | 0.289 | 0.690 | 0.192 | 67 |
| | baseline model + inflammatory exposures | 0.016 | -0.236 | 0.267 | 0.901 | 0.192 | 63 |
| | adjusted model + inflammatory exposures | 0.030 | -0.210 | 0.269 | 0.804 | 0.326 | 63 |
| **VEGFA** | baseline model | -0.014 | -0.261 | 0.234 | 0.913 | 0.190 | 67 |
| | baseline model + inflammatory exposures | -0.033 | -0.286 | 0.221 | 0.797 | 0.193 | 63 |
| | adjusted model + inflammatory exposures | -0.028 | -0.265 | 0.208 | 0.810 | 0.326 | 63 |
| **WFIKKN2** | baseline model | 0.141 | -0.099 | 0.381 | 0.245 | 0.208 | 67 |
| | baseline model + inflammatory exposures | 0.131 | -0.124 | 0.386 | 0.307 | 0.207 | 63 |
| | adjusted model + inflammatory exposures | 0.088 | -0.149 | 0.324 | 0.461 | 0.332 | 63 |
